# Discovery of the sixth *Candida auris* clade in Singapore

**DOI:** 10.1101/2023.08.01.23293435

**Authors:** Chayaporn Suphavilai, Karrie Kwan Ki Ko, Kar Mun Lim, Mei Gie Tan, Patipan Boonsimma, Joash Jun Keat Chu, Sui Sin Goh, Prevena Rajandran, Lai Chee Lee, Kwee Yuen Tan, Bushra Binte Shaik Ismail, May Kyawt Aung, Yong Yang, Jean Xiang Ying Sim, Indumathi Venkatachalam, Benjamin Pei Zhi Cherng, Bram Spruijtenburg, Kian Sing Chan, Lynette Lin Ean Oon, Ai Ling Tan, Yen Ee Tan, Limin Wijaya, Ban Hock Tan, Moi Lin Ling, Tse Hsien Koh, Jacques F. Meis, Clement Kin Ming Tsui, Niranjan Nagarajan

## Abstract

**Background:** The emerging fungal pathogen *Candida auris* poses a serious threat to global public health due to its worldwide distribution, multidrug-resistance, high transmissibility, propensity to cause outbreaks and high mortality rates. We report three *C. auris* isolates detected in Singapore, which are genetically distinct from all known clades (I-V) and represent a new clade (Clade VI).

**Methods:** Three epidemiologically unlinked clinical isolates belonging to the potential new *C. auris* clade were whole-genome sequenced and phenotypically characterized. The complete genomes of these isolates were compared to representative genomes of all known clades. To provide a global context, 3,651 international whole-genome sequences (WGS) from the NCBI database were included in the high-resolution single nucleotide polymorphism (SNP) analysis. Antifungal resistance genes, mating type locus, and chromosomal rearrangements were characterized from the WGS data of the Clade VI isolates. We further implemented Bayesian logistic regression models to simulate the automatic detection of Clade V and VI as their WGS data became available.

**Findings:** The three Clade VI isolates were separated by >36,000 SNPs from all existing *C. auris* clades. These isolates had opposite mating type allele and different chromosomal rearrangements when compared to their closest Clade IV relatives. As a proof-of-concept, our classification model was able to flag these outlier genomes as a potential new clade. Furthermore, an independent WGS submission from Bangladesh was found to belong to this new clade.

**Interpretation:** The discovery of a new *C. auris* clade in Singapore and Bangladesh, showing close relationship to Clade IV members in South America, highlights the unknown genetic diversity and origin of *C. auris*, particularly in under-resourced regions. Active surveillance in clinical settings, along with effective sequencing strategies and downstream analysis, will be essential in the identification of novel strains, tracking of transmission, and containment of adverse clinical impacts caused by *C. auris* infections.

**Funding:** This work was supported by the Singapore National Medical Research Council (NMRC) research training fellowship (MOH-FLWSHP19may-0005), the NCRS Duke-NUS Academic Medical Center Academic Clinical Program grant (09/FY2022/P1/17-A32, GRDUKP003401), and the Genedant-GIS Innovation Program grant.

**Research in context:** *Evidence before this study:* We searched PubMed using the search terms “*Candida auris*” AND “clade”, for papers published between Jan 1, 2009, and July 1, 2023. This search retrieved 115 publications. 60 relevant publications were identified. 28 studies analyzed and discussed the molecular epidemiology of *C. auris*, including the description of *C. auris* clades, either in outbreak or surveillance settings. There were 11 case reports of *C. auris* clinical cases that included clade determination. Two studies focused on the detection and clade determination of *C. auris* from non-healthcare environments. Clade-specific characteristics were described or analyzed in 14 studies. One study applied machine learning to *C. auris* drug resistance analysis, but not for clade determination. Four studies focused on the description of potentially new *C. auris* lineages, subclades, or clades. All publications described isolates that belong to one of the five known *C. auris* clades (I-V). All publications found that strains from different clades differed by more than 35,000 SNPs, and that there are clade-specific differences in geographical distribution, phenotypic characteristics, antifungal susceptibility profile, outbreak potential, and clinical manifestations. The NCBI Pathogen Detection system contained 4,506 *C. auris* genomes on July 1, 2023. There were ten (0·22%) submissions from Southeast Asian countries and 92 (2·04%) submissions from South Asia and the Indian subcontinent, which are parts of the Indomalayan biogeographic realm.

*Added value of this study:* To the best of our knowledge, we are the first group to perform hybrid assemblies on three representative isolates in a new *C. auris* clade, which is separated from all other existing clades (I-V) by >36,000 SNPs. Whole-genome SNP analysis and phenotypic characterization of these epidemiologically unlinked isolates detected in Singapore suggest that they represent a previously unreported sixth major clade. High-resolution SNP analysis of 3,651 international whole-genome sequences from the NCBI database, which generated the final dataset consisting of more than 6.6 million genome pairs, revealed six distinct genetic clusters representing the five known clades and the new sixth clade (Indomalayan). In addition, we demonstrate that a machine learning approach can be used to flag these outlier genomes for further investigations as soon as they become available, thus providing the earliest possible alert for potential new public health threats.

*Implications of all the available evidence:* Despite the high antimicrobial resistance burden in Southeast Asia and South Asia, these regions are disproportionately underrepresented in terms of genomic surveillance of *C. auris*, a multidrug-resistant fungal pathogen. The detection of three epidemiologically unlinked *C. auris* isolates in Singapore belonging to a new *C. auris* clade suggests that yet-to-be-reported strains may be circulating in the region. Given the propensity for multidrug resistance, healthcare-associated infection outbreaks, and the associated high mortality, active surveillance and continued vigilance is necessary.

## Introduction

*Candida auris* is an emerging multidrug-resistant (MDR) fungal pathogen that has been implicated in a large number of healthcare-associated infection outbreaks^1–8^. Invasive infections caused by MDR *C. auris* are associated with high mortality rates in vulnerable patient groups^2–10^. Within healthcare settings, *C. auris* appears to be highly transmissible and is known to persist in the environment^4–6,11^. Given its worldwide distribution, MDR characteristics, high transmissibility, and high mortality rates, *C. auris* represents a serious threat to global public health. The World Health Organization highlighted this threat by classifying *C. auris* as a ‘critical priority’ pathogen last year^12^, while the U.S. Centers for Disease Control and Prevention declared *C. auris* as an ‘urgent’ antimicrobial resistance threat’^13^, further emphasizing the need to understand and mitigate this public health threat.

Five geographically distinct *C. auris* clades have been described to date. This includes Clade I (South Asian), Clade II (East Asian), Clade III (African), Clade IV (South American), and Clade V (Iranian)^10,14–16^. These clades were recognized based on whole-genome sequence analyses, where strains from different clades differed by more than 35,000 single nucleotide polymorphisms (SNPs)^14,17^. Furthermore, these clades differed in terms of their antifungal susceptibility profiles, outbreak potential, and clinical manifestations^18–20^.

Here we report three epidemiologically unlinked *C. auris* isolates detected in Singapore, which appear to represent members of a new clade. The index isolate (A) was first detected by polymerase chain reaction (PCR) from a routine *C. auris* screening swab and subsequently cultured in April 2023. WGS analysis of isolate A showed that the strain was separated from all known clades by more than 36,000 SNPs. Retrospective analysis of our laboratory’s archived *C. auris* isolates subsequently discovered two additional isolates (B and C) with similar genomic and phenotypic characteristics. Isolate C was previously short-read sequenced and reported in 2019 (SRR10102336)^21^.

To understand the global context, we analyzed 3,651 *C. auris* whole-genome sequences in the NCBI database together with the sequence data generated in this study. The final dataset consisted of >6·6 million genome pairs, and six distinct genetic clusters were recognized, consisting of the five known clades and the new Indomalayan clade. From the NCBI database, we identified one additional independently submitted genome from Bangladesh (SRR24877249), which belongs to this new clade. The previously submitted WGS data of isolate C from Singapore (SRR10102336)^21^, and the independent upload from Bangladesh (SRR24877249) were neither identified nor reported as representatives of a potential new clade at the time of writing. The presence of potentially novel, but unrecognized genome data present in the NCBI database demonstrates a gap in novel pathogen or genotype detection despite the availability and accessibility of genomic surveillance data. Given the vast body of microbial genomic data available in the post-COVID-19 pandemic era, it is evident that the challenge lies not only in data availability but also in the ability to fully exploit the data for timely public health alerts and decision-making. To address this gap, we implemented a machine learning approach to demonstrate that potentially novel genomes can be flagged automatically for further investigations, as soon as the genome data become available.

This study reports the hybrid assemblies, genomic analysis, and phenotypic characteristics of three representatives of the sixth *C. auris* clade. We further simulated the automatic detection of the new clade, demonstrating that microbial genomic surveillance can be further augmented to provide early alerts for potential emergent threats.

## Methods

### *Candida auris* microbiological and epidemiological analysis

Three *C. auris* isolates from three epidemiologically unlinked patients in Singapore were included in this study. The full description of the isolates is summarized in appendix 1 (p3). Details of phenotypic identification and antifungal susceptibility testing are in appendix 1 (p4). Four ATCC *C. auris* isolates from Clade I-IV were included as clade-specific controls in this study (appendix 2). The sample and patient identifiers used throughout the manuscript have been modified from the original identifiers, and the identifiers used in the manuscript and supplementary materials cannot reveal the identity of the subjects.

This study used pre-existing retrospective collections of isolates and our analyses led to no clinical intervention. Epidemiological data collection was previously performed as part of routine surveillance and infection prevention measures and hence constituted a non-research infection control surveillance activity. Institutional review board exemption was granted by the SingHealth Centralised Institutional Review Board (Reference number 2017-2576).

### DNA extraction, WGS and sequence data analysis

Details of DNA extraction and WGS methods are in appendix 1 (p4). Methods used for sequence data analysis, including sequence read quality control, genome assembly, variant calling, genome relatedness analysis, phylogenetic analysis, antibiotic resistance gene analysis, mating type loci analysis, chromosomal rearrangement analysis, *in silico* short tandem repeat (STR) analysis, and divergence time estimation are available in appendix 1 (pp 4-7). Quality statistics for the sequence data and genome assemblies generated in this study are summarized in appendix 2. In addition to the new sequence data generated in this study, 4,475 publicly available genomes were initially included, of which 3,651 passed quality control and were included in downstream analysis. The list of publicly available genomes included, and their quality statistics are in appendix 3.

### Machine-learning models for detecting potential new clade

Current methods of clade determination rely on phylogenetic analysis or STR genotyping, the interpretation of which can be challenging, resulting in clade assignments that may be uncertain, as evidenced by the previous report on Isolate C, which was labeled as part of the Clade IV phylogenetic cluster^21^. There is currently no established threshold for SNP distance to flag an unusual *C. auris* genome as a potential new clade. To address this, we implemented a Bayesian logistic regression model to determine whether a pair of genomes are from the same clade based on SNP distances and previously reported clades. This threshold was then used to determine the relationship between genomes (an edge links genomes from the same clade) in a graph, which captured clusters (connected components) representing existing and potential new clades. We implemented the model using 3,651 publicly available whole-genome sequences, and the Clade VI whole-genome sequences generated in this study (appendix 3). 1,132 (31·0%) of the publicly available sequence data had previously reported clade information. All WGS data were grouped into 19 quarterly datasets based on their date of public release (2019 - June 2023). The number of clusters in the graph captured the number of clades present at each time point. Detailed methods regarding the algorithm and model evaluation are in appendix 1(pp 8-11).

## Results

All three Clade VI isolates (A, B, C) were phenotypically identified to be *C. auris*. Details for phenotypic identification results are in appendix 1 (p13). The colony morphology of all Clade VI isolates and the lactophenol cotton blue slide mount of isolate A at 600× magnification are in appendix 1 (p16). Antifungal susceptibility testing results are in **Table 1**. Further results and discussions on antifungal resistance testing are in appendix 1 (pp 22-24).

**Table 1.**
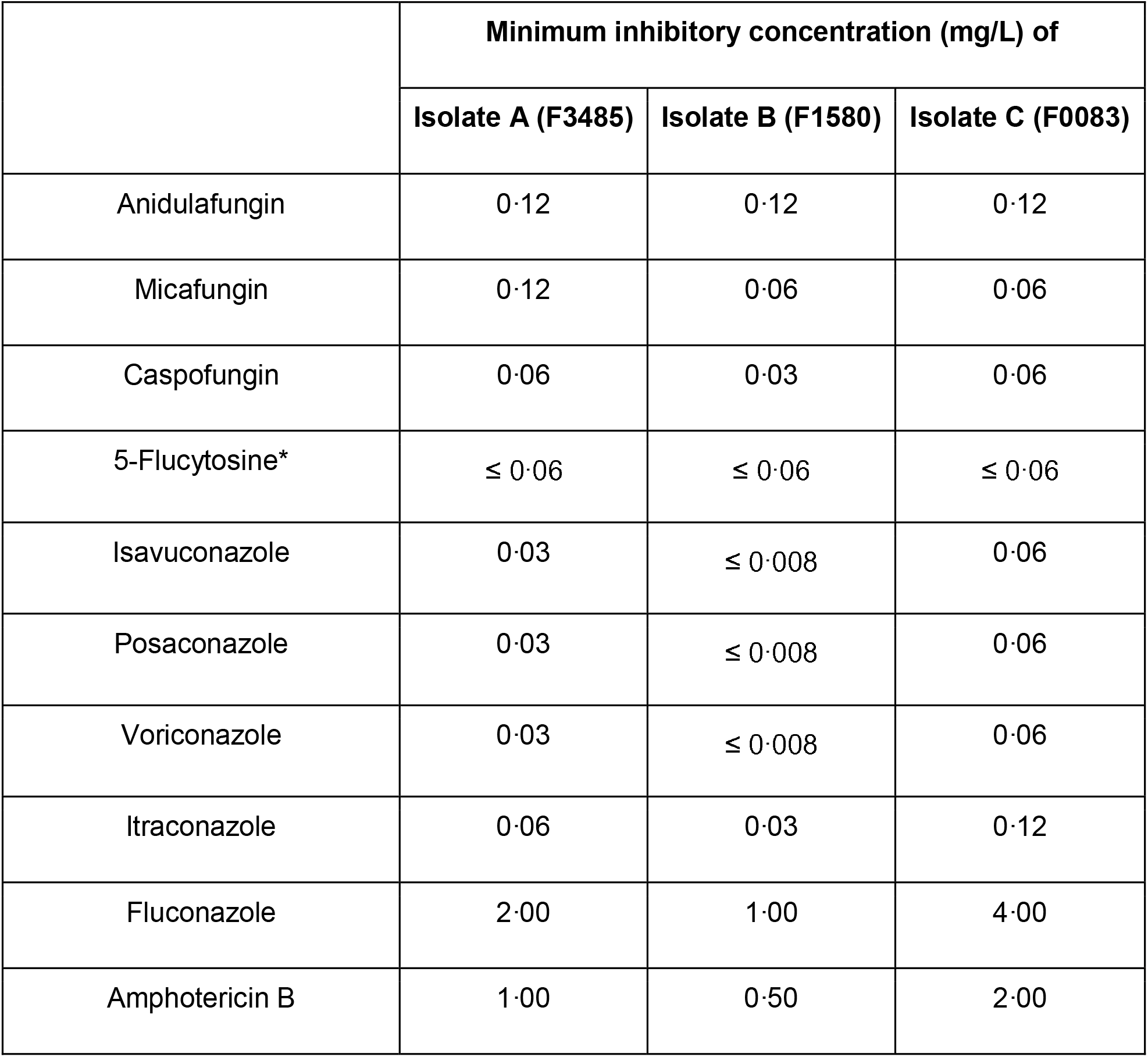
Minimum inhibitory concentration (MIC) values of Singapore Clade VI Candida auris isolates. Antifungal susceptibility testing was performed using the Sensititre™ YeastOne™ ITAMYUCC Plate (Thermo Scientific, Cleveland, OH, USA) according to the manufacturer’s instructions, in a College of American Pathologists-accredited clinical laboratory. *5-Flucytosin MIC was determined using the Sensititre™ YeastOne™ YO10 Plate (International version) (Thermo Scientific, Cleveland, OH, USA). Isolates A and B exhibited wild-type MICs for all tested antifungals^40,41^.

|  | Minimum inhibitory concentration (mg/L) of |  |  |
| --- | --- | --- | --- |
|  | Isolate A (F3485) | Isolate B (F1580) | Isolate C (F0083) |
| Anidulafungin | 0.12 | 0.12 | 0.12 |
| Micafungin | 0.12 | 0.06 | 0.06 |
| Caspofungin | 0.06 | 0.03 | 0.06 |
| 5-Flucytosine* | ≤ 0.06 | ≤ 0.06 | ≤ 0.06 |
| Isavuconazole | 0.03 | ≤ 0.008 | 0.06 |
| Posaconazole | 0.03 | ≤ 0.008 | 0.06 |
| Voriconazole | 0.03 | ≤ 0.008 | 0.06 |
| Itraconazole | 0.06 | 0.03 | 0.12 |
| Fluconazole | 2.00 | 1.00 | 4.00 |
| Amphotericin B | 1.00 | 0.50 | 2.00 |

The index isolate from Patient A, isolate A (LIMS ID F3485, Biosample SAMN36753178) was cultured from a combined screening swab (nasal, axillae, and groin) in April 2023. The screening swab was taken within 24 hours of Patient A’s admission as part of a *C. auris* active surveillance program in the hospital. Patient A received treatment in the Department of Internal Medicine and reported no travel history in the preceding two years. Since *C. auris* is not known to be endemic in Singapore, epidemiological, microbiological, and molecular investigations were initiated. Retrospective analysis of our laboratory’s archived *C. auris* isolates found two additional isolates (B and C) to be similarly genetically distant to the five known clades, out of 43 isolates analyzed. Isolate B (LIMS ID F1580, Biosample SAMN36753179) was cultured from an intraoperative tissue specimen from Patient B, in April 2022. Patient B received treatment in the orthopedic unit and reported no recent travel history in the preceding two months, but had outpatient visits at hospitals in Indonesia in 2021. Isolate C (LIMS ID F0083, Biosample SAMN36753180) was cultured from the positive blood culture of Patient C in January 2018.

Patient C was admitted directly from Bangladesh and the blood culture that was taken within 48 hours of admission grew *C. auris*. Isolate C was previously sequenced in 2019 and is one of the ten publicly available *C. auris* genomes submitted from Singapore (SRR10102336)^21^.

Extensive epidemiological investigations determined that Patients A, B, and C were separated spatially and temporally, with no overlap in physical space, clinical unit, and care facilities utilization in the large 1700-bed tertiary hospital. In accordance with the hospital’s infection prevention policy, every positive case of *C. auris* triggered epidemiological investigations and contact tracing. All inpatients who shared the same ward or room with the positive cases were defined as contacts and they were screened for *C. auris* colonization as part of the infection prevention investigations. A total of 68 contacts were identified for Patients A and B. Patient C was isolated on admission and had no known contacts. None of the contacts screened positive for *C. auris* by routine diagnostic *C. auris* in-house validated PCR test or cultures. Furthermore, a total of 51 environmental samples were cultured to look for *C. auris* during the investigations, none of which yielded *C. auris*.

All three Clade VI *C. auris* isolates were whole-genome sequenced using two sequencing platforms: Illumina (short-read technology) and Oxford Nanopore (long-read technology), to obtain high-quality hybrid assemblies of the genomes. *De novo* hybrid genome assembly of the *C. auris* genomes was performed for the Singapore Clade VI isolates, and each consisted of eight to nine contigs, corresponding to near-complete genomes. The total assembly size was 12·4 to 12·5 Mbp. Quality statistics for Clade VI isolate sequence data and assemblies are available in appendix 2.

A comprehensive genome relatedness analysis using 3,651 *C. auris* WGS data in the NCBI database and sequence data generated in this study was performed (appendix 3). The final dataset included a total of 3,654 whole-genome sequences after quality control checks (appendix 1 p5), and >6·6 million genome pairs were analyzed. We found six distinct genetic clusters in this large-scale analysis. One additional *C. auris* genome (SRR24877249), independently submitted from Bangladesh, was found to belong to the same cluster as the Singaporean Clade VI isolates.

Since complete chromosomal sequences allow us to perform both whole genome phylogenetic and chromosomal rearrangement analyses, 14 representative genomes from known clades (I-V) with complete sequences of chromosomes were selected for in-depth downstream analysis. Phylogenetic analysis of the Clade VI genomes with these representative genomes showed that the Clade VI genomes were genetically distinct from the five known clades (**Figure 1A**). The four newly assembled Clade VI genomes clustered together in the new clade, which was more closely related to Clade IV but differed from all five known clades by at least 36,000 SNPs. The SNP distances between clades are shown in **Figure 1B**. The number of SNP differences within each clade is in appendix 1 (p17). *In silico* STR analysis was performed based on a previously published scheme, and the topology of the dendrogram also demonstrated clustering of Clade VI isolates based on STR profiles, congruent with the observation of a sixth clade using WGS ^22,23^ (appendix 1 p18). Additional Bayesian phylogenetic analysis suggests that the estimated divergence time of Clade VI from the most closely related Clade IV was at 4831 BCE (HPD 272·4 – 311·6 years ago). Details of the molecular clock analysis are in appendix 1 (p19).

**Figure 1.**
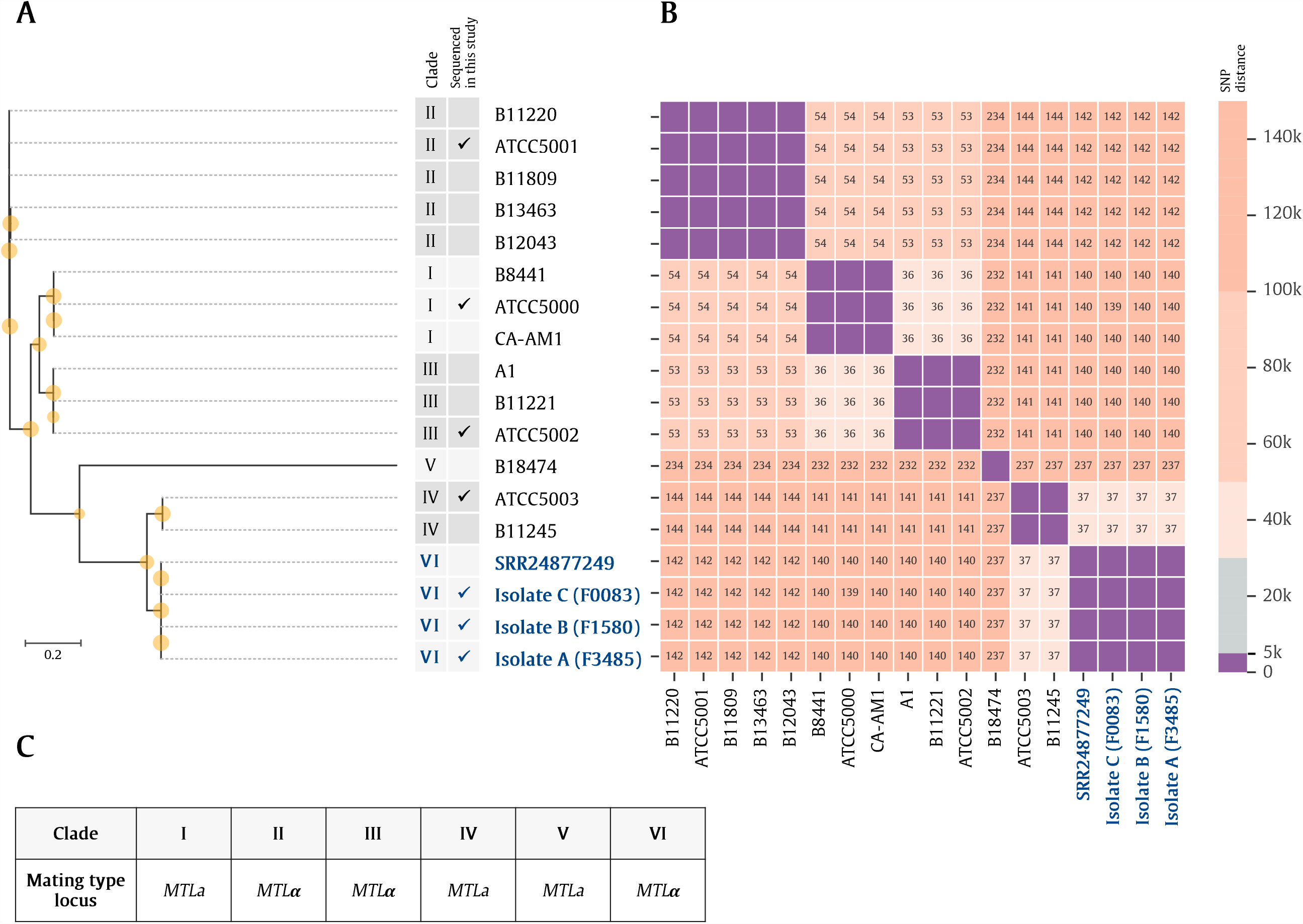
(A) Genetic relationships among representative *Candida auris* genomes from five known clades and the newly identified sixth clade. Six distinct genetic clusters were observed, each consisting of genomes from the same clade. Genetic relationships were inferred using maximum-likelihood phylogenetic analysis, with 1,000 bootstrap replicates. The publicly available B8441 genome was used as a reference to identify small nucleotide polymorphisms (SNPs) in other isolates. Hybrid assemblies of the Singaporean *C. auris* isolates A (F3485), B (F1580), and C (F0083), were used in this analysis. An additional publicly available whole-genome sequence independently submitted from Bangladesh (SRR24877249) was found to belong to the same cluster as the Singaporean Clade VI isolates. The Clade VI genomes are labeled in blue bolded fonts. Yellow circle denotes the bootstrap value. The scale bar represents the mean number of nucleotide substitutions per site. (B) SNP difference among representative C. auris genomes. The numbers within each box indicate the number of SNP differences observed between genomes in distinct clades (in thousands). Isolates between different clades were divergent in at least 35,000 positions genome-wide (e.g. between Clades I and III). Clade VI genomes were divergent in at least 36,000 SNPs from all known clades (I-V). Purple boxes within the heatmap represent genomes that are divergent in ≤5,000 SNPs from each other. (C) Summary of mating type predictions for each clade, determined based on the MTLa/α alleles present at the MAT locus.

We further investigated whether the Clade VI isolates have clade-specific features that distinguish them from their closest relatives in Clade IV. Previous studies observed clade-specific chromosomal rearrangements in *C. auris*^24,25^. We reconstructed complete genomes to investigate if similar clade-specific large chromosomal rearrangements exist in the Singaporean Clade VI isolates (**Figure 2** and appendix 1 p20). Compared to Clade IV, an inversion in chromosome 5 was not observed in the Singaporean Clade VI genomes relative to Clade I. Furthermore, even though the new Clade VI isolates were most closely related to Clade IV, we found that they were of opposite mating types (**Figure 1C**). Clade IV isolates carry the *MTLa* allele at the *MAT* locus^24^, while all Clade VI isolates belong to mating type α with amino acid identity of homologs above 95% based on BLAST matches (appendix 1 p21).

**Figure 2.**
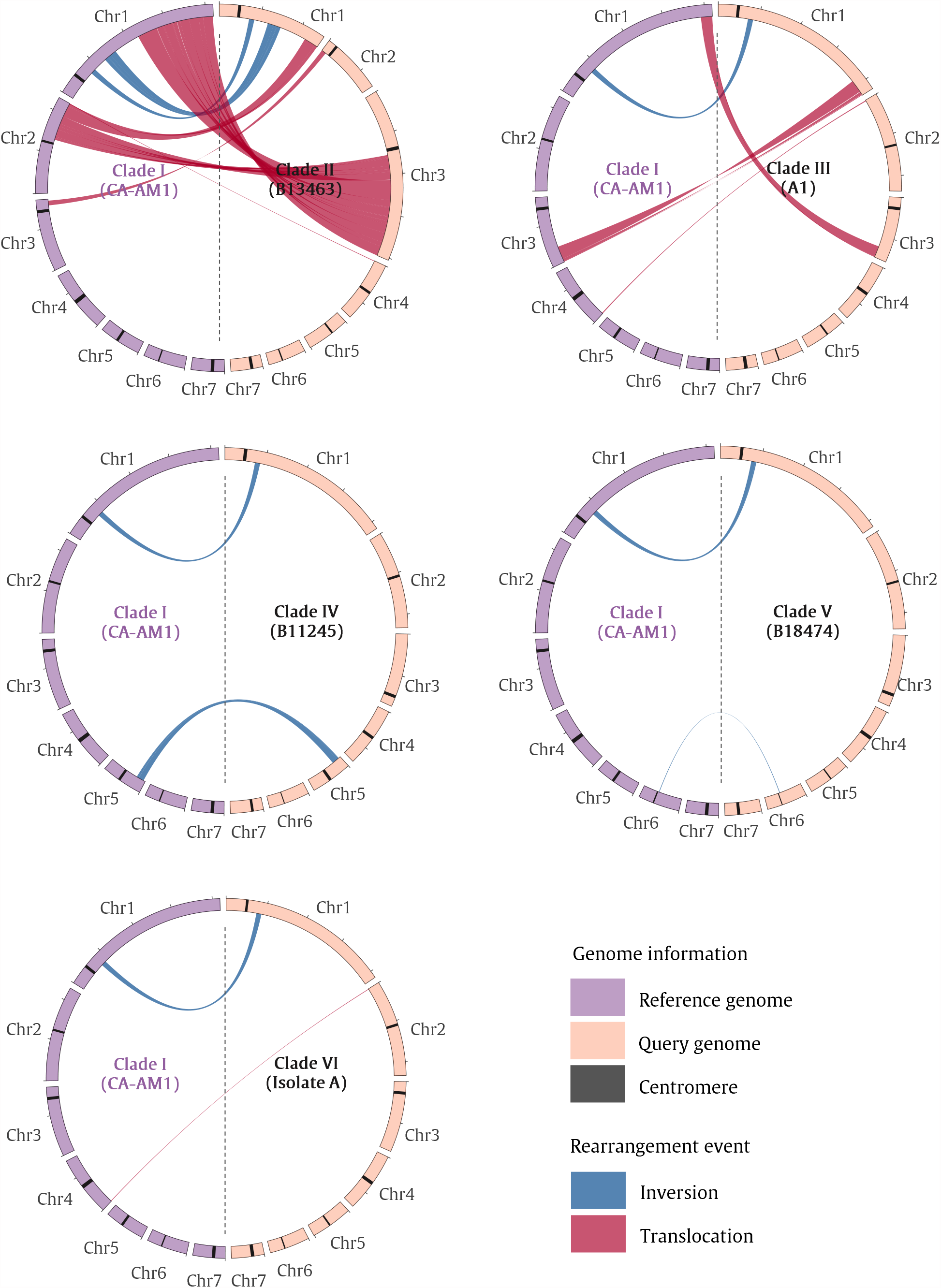
Chromosomal rearrangements in *Candida auris*. Chord diagrams showing translocations and inversions in representative genomes for clades II, III, IV, V, and VI, using CA-AM1 (Clade I) as a reference. Only rearrangement events of size >10,000bp are displayed. Based on whole-genome alignments, Clade VI has an inversion in the long arm of chromosome 1, which is also observed in the representative genomes from Clades II, III, IV, and V. We also identified a unique translocation event between chromosomes 2 and 4 in Clade VI. Representative isolate genomes used in this figure: Clade I, CA-AM1 (GCA_014673535); Clade II, B13463 (GCA_016495665); Clade III, A1 (GCA_014217455); Clade IV, B11245 (GCA_008275145); Clade V, B18474 (SRR13269545); Clade VI, isolate A (F3485).

Given the marked difference among whole genomes of *C. auris* from different clades, we exploited the SNP distances between the genomes to develop a system to monitor and alert for the emergence of potentially new clades. Using publicly available WGS data of 3,651 isolates and the three representative genomes from Clade VI, with a final dataset of >6·6 million genome pairs, we simulated the detection of Clade V and Clade VI at two time points, when the WGS data became available (**Figure 3**). Although the amount of WGS data increased from 554 genomes in 2019 Q1 to 3,654 genomes in 2023 Q3 (appendix 1 pp25-26), the SNP distance thresholds inferred by the Bayesian logistic regression model were consistently between 19,000 – 23,000 SNPs across the time points (**Figure 3A)**. The model detected the emergence of Clades V (2019 Q3) and VI (2023 Q2) as it monitored the SNP distance amongst the genomes over time (**Figure 3B)**. As an illustrative example, in 2023 Q3, there were six connected components and each component consisted of genomes from the same clade (**Figure 3C**). Detailed results on the automatic detection of new clades can be found in appendix 1 pp 25-26.

**Figure 3.**
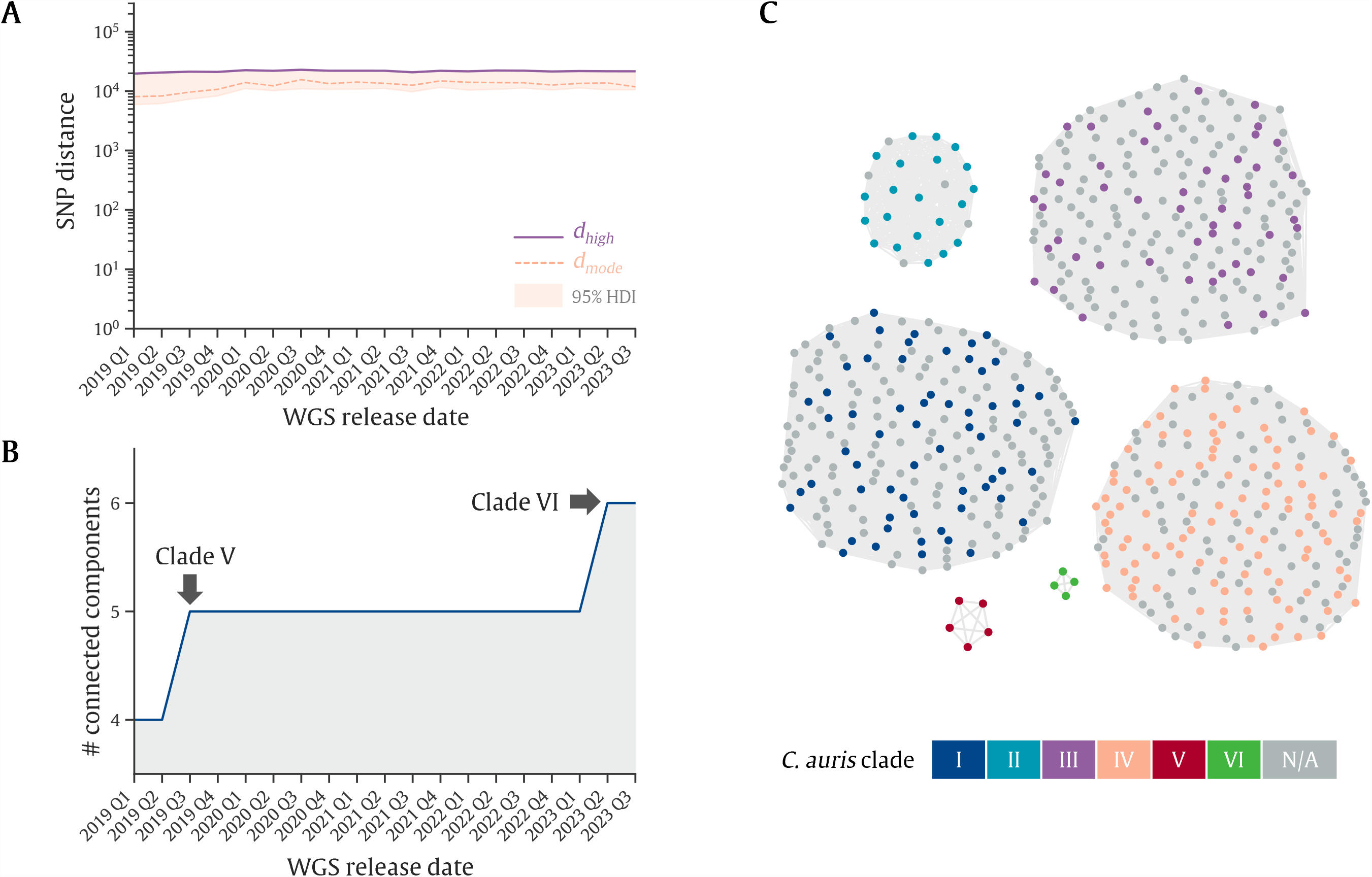
Detection of potentially new *Candida auris* clades using a Bayesian analysis framework. **(A)** Line plot illustrates small nucleotide polymorphism (SNP) distance thresholds learned across time points using Bayesian logistic regression. The Y-axis represents the SNP distance between genome pairs. The pink dashed line represents the mode of the SNP distance threshold (d), and the shaded region represents the 95% high-density interval (HDI) bounded by *d*_*low*_ and *d*_*high*_. The purple solid line marks *d*_*high*_. When the SNP distance is greater than *d*_*high*_, the input genome pair is predicted to be from different clades. The X-axis represents time in quarterly units, where *C. auris* whole genome sequence (WGS) data released within the same time period were added incrementally into the training dataset. **(B)** Line plot illustrates the number of connected components (Y-axis) detected over time (X-axis in quarterly units). The emergence of Clade V and Clade VI were detected by the rise in the number of connected components (CCs) (black arrows). **(C)** Graph visualization of the relationships between genomes in 2023 Q3. Each node represents a *C. auris* genome, and node color denotes clades. Each color represents genomes with known clade information. Gray nodes represent genomes without clade information in the literature. For ease of visualization, we subsampled the genomes in the components which contain more than 200 genomes. An edge links two nodes when *s* < *d*_*high*_, indicating that the genomes are predicted to belong to the same clade. Six connected components were observed across 3,654 genomes.

## Discussion

We report the discovery of the sixth major clade of *C. auris*, which includes three epidemiologically unlinked isolates detected in Singapore, and one publicly available WGS data of an isolate reported from Bangladesh (SRR24877249). This conclusion is based upon extensive genomic analysis demonstrating the clustering of these four isolates, and large genetic divergence from the five known clades. We obtained high-quality hybrid genome assemblies of the Singaporean Clade VI isolates, and analyzed these genomes together with 3,651 publicly available *C. auris* whole-genome sequences.

Although the samples that yielded these isolates were collected in Singapore and Bangladesh, the Clade VI isolates were most closely related to representatives of Clade IV (divergence of ∼36,000 SNPs), which were mostly from South America. The genomic distance observed between Clades IV and VI is greater than that observed between Clades I and III (∼35,000 SNPs). Within Clade VI, the three Singaporean isolates were distinct in at least 70 SNPs. This within-clade difference is similar to what was previously reported among Iranian Clade V isolates, which were cultured from patients with community-onset otomycosis^15^. The independent detection of Clade VI isolates from Singapore and Bangladesh suggests that Clade VI strains might have been circulating undetected in Southeast Asia and South Asia. Given the estimated divergence time from Clade IV, we speculate that there may exist an endemic zone or a special ecological habitat for Clade VI *C. auris* in the Indomalaya biogeographic realm. Further discussion on the divergence time is available in appendix 1 p25.

The Clade VI isolates in Singapore were distinct from their closest relatives in Clade IV based on phenotypic investigations and genomic analysis. Firstly, the susceptibility to antifungals, and lack of known AMR resistance-conferring mutations in the Clade VI isolates (**Table 1** and appendix 1 p22-24) are unlike that of Clade IV, where approximately half of the isolates are resistant to fluconazole^10,14,26,27^. Secondly, Clade VI isolates lack an inversion observed in Clade IV (**Figure 2** and appendix p20). Thirdly, Clade VI isolates carry the *MTL*α allele, a mating-type allele opposite to the *MTLa* observed in Clade IV. As isolates of opposite mating types are yet to be found within the same clade, taken together with other findings presented in this study, the Clade VI isolates are likely representatives of a new major clade rather than a sub-lineage of an existing clade. Further discussion on mating type can be found in appendix 1 p25.

Unlike the five known *C. auris* clades that are named after their geographic location, these Clade VI isolates detected in Singapore have no known specific geographic associations yet. Due to the retrospective nature of this study, extensive travel history could not be obtained from the patients. We postulate that isolate C was related to Bangladesh geographically, as Patient C was airlifted from Bangladesh to Singapore and isolate C was grown from the blood culture taken within 48 hours of his admission. The independent detection of another Clade VI isolate in Bangladesh (SRR24877249, sample collected in 2021) suggests that Clade VI strains may be circulating undetected in Bangladesh, with the first documented Clade VI isolate being detected in Singapore (isolate C, F0083). Patient B was known to have epidemiological links to Indonesia and Singapore, but not to Bangladesh or South Asia. Patient A was not known to have left Singapore in the preceding two years and has no known epidemiological link to South Asia. The identification of more Clade VI isolates may help us to pinpoint a geographic origin for this emergent clade. Further discussion on the detection and containment of *C. auris* in Singapore is in appendix 1 p25.

While the origin of Clade VI cannot be resolved, the cryptic circulation of Clade VI *C. auris* in Southeast Asian and South Asian countries is evident because all four patients had epidemiological links to Bangladesh, Indonesia, and Singapore. Due to the paucity of *C. auris* data in the region, it is unclear whether Clade VI is widespread or expanding from its presumed endemic zone. Its potential for causing outbreaks also remains unknown. Clades I, III, and IV are known to cause prolonged outbreaks in healthcare settings^3,6,14,27–31^. As a result, the acquisition of Clades I, III, and IV *C. auris* is often traced to healthcare settings. On the contrary, Clades II and V are not known to be healthcare-associated^10,15,32,33^. Although the generally wild-type antifungal susceptibility profile of Clade VI resembles that of Clades II and V, two of the three Clade VI isolates were detected in invasive clinical infections (blood and intraoperative tissue). This is unlike Clades II and V, which are most commonly associated with community-onset otomycosis^10,15,32,33^. As the virulence and outbreak potential of Clade VI *C. auris* is yet to be established, it is imperative to ensure its early detection and containment for the purpose of patient safety. Since we have yet to identify the source, the risk factor(s) for acquisition, and the specific geographical associations of Clade VI *C. auris*, active surveillance strategies and priorities may need to be reassessed by individual healthcare institutions, to balance possible emergent threat containment with optimal healthcare resource utilization.

The Clade VI isolates were detected as part of a microbial genomic surveillance program within a Singaporean academic medical center. Although laboratories are increasingly able to obtain microbial genome sequences, it is evident that ongoing, large-scale genomic analysis to provide relevant public health alerts continues to be challenging. We aimed to augment our ability to detect outlier genomes, such as the Clade VI genomes reported in this study, by demonstrating a proof-of-concept machine learning approach that can automatically identify outlier genomes (detection of Clades V and VI) for enhancing genomic surveillance (**Figure 3**). Further discussion on the results and limitations of this machine learning approach can be found in appendix 1 pp25-26.

This study has several limitations. Due to the retrospective nature of this study, full travel and exposure history could not be traced. The sample size of Clade VI isolates was small, and therefore the reported genomes may not be representative of the entire clade. The antifungal susceptibility test was not performed with a reference broth dilution method but was performed with Sensititre™ YeastOne™ plates. Stringent quality control of publicly available WGS data discarded 18.4% (824/4475) of available genome data, including isolate C’s sequence (SRR10102336) reported from Singapore in 2019^21^. This significant data loss from suboptimal quality suggests that when applied in practice, our mathematical modelling approach to detect new clades will face additional challenges, and the framework presented in this study therefore represents a proof-of-concept that requires further evaluations and fine-tuning.

Worsening spread of *C. auris* has recently been reported in Europe and in the United States^34–36^. It is evident that *C. auris* continues to be a threat to public health notwithstanding the resources available for surveillance and containment in high-income countries. Given the resource limitation in parts of Indomalaya, innovative solutions are necessary to augment regional microbial genomic surveillance. Cost-effective integration of existing routine infectious disease diagnostics with genomic surveillance at sentinel sites^37–39^, together with effective sequence data analysis, are essential components for pandemic preparedness. Our study provides evidence for the discovery of the sixth major *C. auris* clade in Singapore and underscores the need to enhance genomic surveillance in strategic sentinel sites.

## Data Availability

Reads and genome assemblies from this study have been deposited in the National Centre for Biotechnology Information (NCBI) Sequence Read Archive (SRA) database (https://www.ncbi.nlm.nih.gov/sra) under BioProject accession number PRJNA1000034. All data produced in the present study are available upon reasonable request to the authors.

## Contributors

CS and KK conceptualized the study and CS, KK, and CT prepared the draft manuscript. CS, KK, CT, KML, and MGT performed the literature search. KML, CT, and NN contributed to the conceptualization and design of the study. MGT, ALT, and YET contributed isolates for analyses and provided expert advice on clinical mycology. CS, KK, KML, PB, SSG, PR, BS and JJKC contributed to the data acquisition and analysis process. KSC and LO contributed samples for analysis and provided expert advice on clinical molecular microbiology. LCL, MLL, BBSI, MKA, YY, JXYS, and IV provided epidemiological data and expert infection control and prevention advice. BC and LW provided specialist infectious disease inputs and clinical data. JFM provided expert advice and supported STR and phylogenetic timing analysis. BS performed STR and phylogenetic timing analysis. BHT and THK critically evaluated the clinical, microbiological, and epidemiological data and analysis. NN critically evaluated the genomic data analysis. All authors contributed to the manuscript, and edited and reviewed the final manuscript. CS and KK accessed and verified the raw data. CS and KK contributed equally to this work. All authors had full access to all the data in the study and had final responsibility for the decision to submit for publication.

## Data sharing

The code used to perform WGS analyses and train machine-learning models in this study is available at https://github.com/CSB5/Candida_auris_CladeVI. Reads and genome assemblies from this study have been deposited in the National Centre for Biotechnology Information (NCBI) Sequence Read Archive (SRA) database (https://www.ncbi.nlm.nih.gov/sra) under BioProject accession number PRJNA1000034.

## Supplementary Material

Supplementary Appendix 1

Supplementary Appendix 2

Supplementary Appendix 3

## Acknowledgements

This work was supported by the NCRS Duke-NUS Academic Medical Center Academic Clinical Program grant (09/FY2022/P1/17-A32, GRDUKP003401), the Genedant-GIS Innovation Program grant, resources from the Genome Institute of Singapore (GIS), and resources from Singapore General Hospital (SGH). Karrie K. K. Ko is supported by the Singapore National Medical Research Council (NMRC) research training fellowship (MOH-FLWSHP19may-0005). The work was supported in part through computational resources (Sockeye) and services provided by Advanced Research Computing at the University of British Columbia, Vancouver, Canada. We thank members of the Diagnostic Bacteriology Laboratory and the Molecular Laboratory, SGH, for their technical assistance. We thank members of the Department of Infection Prevention and Epidemiology, SGH, for their assistance with epidemiologic data. We thank the Department of Infectious Diseases, SGH, for their assistance with clinical data. We thank members of the Laboratory of Metagenomic Technologies and Microbial Systems, GIS, for their helpful comments on genomic analysis.

