## Supplementary Appendix 1 for "Discovery of the sixth *Candida auris* clade in Singapore"

**Discovery of the sixth *Candida auris* clade in Singapore**

Chayaporn Suphavilai^^1^, Karrie Kwan Ki Ko^*^1,2,3,4,5^, Kar Mun Lim^1^, Mei Gie Tan^2^, Patipan Boonsimma^1^, Joash Jun Keat Chu^1^, Sui Sin Goh^2^, Prevena Rajandran^2^, Lai Chee Lee^6^, Kwee Yuen Tan^6^, Bushra Binte Shaik Ismail^6^, May Kyawt Aung^6^, Yong Yang^4,6^, Jean Xiang Ying Sim^4,6,7^, Indumathi Venkatachalam^4,6,7^, Benjamin Pei Zhi Cherng^4,7^, Bram Spruijtenburg^8,9^, Kian Sing Chan^3,4^, Lynette Lin Ean Oon^3,4^, Ai Ling Tan^2,4^, Yen Ee Tan^2,4^, Limin Wijaya^4,7^, Ban Hock Tan^4,7^, Moi Lin Ling^4,6^, Tse Hsien Koh^2,4^, Jacques F. Meis^8,9,10^, Clement Kin Ming Tsui^11,12,13^, Niranjan Nagarajan*^1,5^

^Contributed equally to this work

*Corresponding authors

1. Genome Institute of Singapore, Agency for Science, Technology and Research (A*STAR), Singapore
2. Department of Microbiology, Singapore General Hospital, Singapore
3. Department of Molecular Pathology, Singapore General Hospital, Singapore
4. Duke-NUS Medical School, Singapore
5. Yong Loo Lin School of Medicine, National University of Singapore, Singapore
6. Department of Infection Prevention and Epidemiology, Singapore General Hospital, Singapore
7. Department of Infectious Diseases, Singapore General Hospital, Singapore
8. Department of Medical Microbiology and Infectious Diseases, Canisius-Wilhelmina Hospital, Nijmegen, The Netherlands
9. Center of Expertise in Mycology Radboud University Medical Center/Canisius-Wilhelmina Hospital, 6532 SZ Nijmegen, The Netherlands
10. Department I of Internal Medicine, University Hospital Cologne, Excellence Center for Medical Mycology, Cologne, Germany
11. Infectious Diseases Research Laboratory, National Centre for Infectious Diseases, Tan Tock Seng Hospital, Singapore
12. Lee Kong Chian School of Medicine, Nanyang Technological University, Singapore
13. Faculty of Medicine, University of British Columbia, Vancouver, BC, Canada

### **Supplementary Table 1**

| **Sample name in manuscript** | **Isolate A** | **Isolate B** | **Isolate C** |
| --- | --- | --- | --- |
| **Patient name** | A | B | C |
| **Manuscript LIMS ID** | F3485 | F1580 | F0083 |
| **Source** | Nasal, axillae, and groin swab | Intraoperative tissue | Blood |
| **Date of clinical sample collection** | 2023 | 2022 | 2018 |
| **Existing clade identified** | None | None | None |
| **First sequenced in this study** | Yes | Yes | No |
| **Multiple isolates from the same patient?** | No | No | No |
| **Nanopore Sequencing Run ID for this study** | N521_BC01 | N466_BC12 | N494_BC21 |
| **Illumina Sequencing Run ID for this study** | MDB1198, MDB1217 | WMB1910, MDB1197 | MDB1218 |
| **Accession prior to this study** | NA | NA | PRJNA540907, Biosample SAMN11866133 |
| **Accession generated in this study** | BioProject PRJNA1000034 Biosample SAMN36753178 | BioProject PRJNA1000034 Biosample SAMN36753179 | BioProject PRJNA1000034 Biosample SAMN36753180 |

**Supplementary Table 1**. Singapore Clade VI isolates metadata

### **Supplementary Methods**

#### **Phenotypic identification and antifungal susceptibility testing**

All isolates were identified by matrix-assisted laser desorption ionization-time of flight (MALDI-TOF) mass spectrometry (BrukerDaltonics, Billerica, MA, United States) using *in-vitro diagnostic* (IVD) library revision K (2020) software following the manufacturer’s instructions. Phenotypic biochemical profiles were obtained using the Vitek2 YST ID Card (bioMérieux, Marcy-l'Étoile, France) and API 20C AUX (bioMérieux, Marcy-l'Étoile, France). Antifungal susceptibility testing was performed using the Sensititre^TM^ YeastOne^TM^ ITAMYUCC and YO10 AST plates (Thermo Scientific, Cleveland, OH, USA) according to the manufacturer’s instructions, in a College of American Pathologists (CAP)-accredited clinical laboratory.

#### **DNA extraction and whole genome sequencing**

DNA was extracted using the Qiagen DNeasy PowerSoil Pro Kit 47014 (Qiagen, Venlo, The Netherlands). Illumina sequencing was performed as per the manufacturer's instruction on either the HiSeq X Five platform or the MiniSeq platform (Illumina, San Diego, CA, United States). Long-read Nanopore sequencing was performed using the Ligation Sequencing Kit SQK-LSK109 (Oxford Nanopore Technologies, Oxford, United Kingdom) and the Native Barcoding Kits EXP-NBD104 and EXP-NBD114 according to the manufacturer's instructions on MinION R9.4.1 FLO-MIN106D flowcells (Oxford Nanopore Technologies) on a Mk1C sequencer. For Nanopore sequencing, the acquisition of the sequenced reads was carried out using MinKNOW v21.11.6. Base-calling, demultiplexing, and trimming of barcodes and adaptor sequences were carried out via the Guppy v5.0.7 Super High Accuracy basecaller (Oxford Nanopore Technologies).

#### **Sequence data analysis**

##### **Overview of whole-genome sequence (WGS) data sequenced in this study**

Seven *Candida auris* isolates were whole-genome sequenced in this study. These include three Clade VI isolates and four control isolates for Clade I-VI (Supplementary Appendix 2). All seven isolates were sequenced using long-read Nanopore sequencing platform. Additional short-read Illumina sequencing was performed for the three Clade VI isolates, and Clades I and IV control isolates.

##### **Overview of WGS data analyzed in this study**

Two types of public WGS data were obtained for this study. Firstly, for the analysis of 18 representative genomes, additional nine publicly available assembled genomes were obtained from National Center for Biotechnology Information (NCBI) Genome Database and the Candida Genome Database (CGD) (Supplementary Appendix 2). An additional long-read nanopore WGS data of Clade V isolate (B18474, SRR13269545) was obtained from NCBI database.

Secondly, for the large-scale analysis and machine learning models, a list of 4,358 Illumina WGS accessions together with data release date information was obtained from NCBI Pathogen Detection (https://www.ncbi.nlm.nih.gov/pathogens) on 29 June 2023. This list was then combined with the curated list of 1,285 *Candida auris* WGS data with clade information reported in the previous studies^1,2^. In total, 4,475 unique Illumina WGS data were included in this study. (Supplementary Appendix 3)

##### **Sequencing analysis pipelines**

Sequencing analysis pipelines were developed for processing Illumina and Nanopore WGS data using Nextflow v23.04.2^3^. The following sections described the key steps implemented within the pipeline, including sequence data preprocessing, genome assembly, and variant calling. The Nextflow pipelines and Docker containers for sequencing analysis and custom scripts for machine learning are available at https://github.com/CSB5/Candida_auris_CladeVI.

##### **Illumina sequencing data preprocessing**

For Illumina paired-end sequencing reads, WGS data were downloaded from NCBI Sequence Read Archive (SRA) database using SRA Toolkit v3.0.5 (https://github.com/ncbi/sra-tools). Adapter was trimmed and reads quality were filtered using fastp v0.23.2^4^ (with Phred quality of Q18, the unqualified bases limit was set to 40%, the average quality score was set to 18 to discard the read, maximum of N bases in a read was set to 5, minimum read length of 100, --low_complexity_filter option was set, and the complexity threshold was set to 30%). Sequencing quality statistics were calculated using FastQC v0.12.1^5^ Read statistics were calculated using SeqKit v2.5.0^6^. After quality filtering, sequencing depth was calculated and WGS data with the depth lower than 30✕ were discarded. The maximum depth was set to 100✕, and WGS data exceeding the maximum depth were subsampled. The pre-processed fastq files were used for genome assembly and other downstream analyses. The detailed statistics are in Supplementary Appendix 3.

##### **Nanopore sequencing data preprocessing**

For Nanopore sequencing reads, read quality was filtered using chopper v0.5.0^7^ (with Phred quality of Q10, and minimum read length of 500). Read statistics were calculated using NanoStat v1.6.0^8^ and SeqKit v2.5.0^6^. After quality filtering, sequencing depth were calculated and WGS data with less than 30✕ depth were discarded. The maximum depth was set to 150✕, and WGS data exceeding the maximum depth were subsampled. The pre-processed fastq files were used for genome assembly and other downstream analyses. The detailed statistics are in Supplementary Appendix 2.

##### **Genome assembly**

###### **Ilumina-based assembly**

*De novo* genome assembly for the paired-end Ilumina sequencing reads was performed using SPAdes v3.15.5^9^ (with --isolate option to improve assembly quality). Contigs that were shorter than 1,000 nucleotides were removed using SeqKit v2.5.0^6^. The detailed statistics, including number of contigs, mean sequencing depth, and total genome size are in Supplementary Appendix 3.

###### **Nanopore-based assembly**

*De novo* assembly of draft genomes for Nanopore reads was performed using Flye v2.9.1^10^ (with --nano-hq option was set to indicate high-quality reads). The draft genomes were then polished by Medaka v1.8.1 (Oxford Nanopore Technologies, Oxford, United Kingdom) with a pre-processed read and a draft sequence to correct a result from Flye. Additional polishing was performed using pre-processed reads, and the consensus sequence from the first round of polishing. Finally, contigs that are shorter than 1,000 nucleotides were filtered out using SeqKit v2.5.0^6^. The detailed statistics, including number of contigs, mean sequencing depth, and total genome size, are in Supplementary Appendix 2.

###### **Hybrid genome assembly**

To further improve the quality of the assembled genomes, genome polishing was performed using Pilon v1.24^11^ (with --frags option was set to use sorted index file in assembly step). Briefly, the nanopore-based assembled genomes were indexed using BWA v0.7.17^12^. The Illumina short reads were aligned to the indexed genome using BWA v0.7.17^12^ and SAMtools v1.14^13^. Finally, the consensus genome was constructed from the long-read genome by using Pilon. The detailed statistics, including number of contigs, mean sequencing depth, and total genome size are in Supplementary Appendix 2.

##### **Species identification**

To build a database for fungal species identification, all 544 fungal genomes were downloaded from Refseq database (as of 29 June 2023). Mash similarity was calculated between the input assembled genomes and each fungal genome in the database (Mash v2.3 with k=21 and s=10000)^14^. A mash distance threshold of 0·04, similar to that of Pathogen Watch’s Speciator^15^ and Kleborate^16^, was used to confirm the species of the input assembled genomes.

Additional species confirmation was performed for the 18 representative genomes by extracting the internal transcribed spacer (ITS) sequence from the assembled genomes by BLASTn v2.9.0^17^. Percent identity of 95% and ITS region coverage of 95% was used for identifying the best-match species. Full-length *Candida auris* ITS sequence (NR_154998.1) was detected in all 18 representative genomes (Supplementary Appendix 2).

##### **Phylogenetic analysis**

Using B8441 (Candida Genome Database) as a reference genome, phylogenetic analysis of the 18 representative genomes was performed using Parsnp v1.2^18^ with option -x to enable filtering of small nucleotide polymorphisms (SNPs) located in regions of recombination and -c to include all genomes. The coverage of 18 genomes were confirmed to be greater than 85% (85.5-91.7%). Harvesttools v1.2^18^ was used for extracting SNP information and creating a vcf file. SNP distance was calculated using snp-dists v0.8.2. The final maximum-likelihood phylogenetic tree was generated using RAxML-NG v1.1.0 (with GTR+G model and 1,000 bootstrap replicates)^19^.

##### **Variant calling**

Variant calling of large-scale analysis of Illumina WGS data was performed using Snippy v4.6.0 (<https://github.com/tseemann/snippy>) with the default parameters. Additionally, we filter only SNPs with an alternative allele frequency of at least 0·9. All output vcf files were then combined for downstream analysis.

As an additional validation of our variant calling results, the numbers of SNPs reported by our Snippy-based pipeline were compared against the recent benchmarking study^20^, consisting of 12 *Candida auris* variant calling pipelines. There were 22 Illumina WGS data matched with the benchmarking study. Using the same B8441 as a reference genome, the numbers of SNPs reported by our pipeline were consistent with the previous study. Briefly, there were <1,500 SNPs for Clade I genomes, >30,000 SNPs for Clade II and Clade III genomes, and >100,000 SNPs for Clade IV genomes.

##### **Genome characterization**

###### **Identifying antifungal-resistance mutations**

To identify antifungal resistance-conferring mutations, we aligned the 18 representative genomes to the reference genomes B8441 (GCA_002759435.2) using NUCleotide MUMmer (NUCmer) v3.1^21^. The vcf files were extracted and annotated using SnpEff v5.1^22^ (with candida_auris_GCA_002759435.2 database). We then inspected specific loci containing genes known to be relevant to AMR in *Candida auris*, including lanosterol 14 α-demethylase (*ERG11*), Sterol 24-C-methyltransferase (*ERG6*), 1,3-beta-glucan synthase (*FKS1*), zinc-cluster transcription factor-encoding gene *TAC1B*, and uracil phosphoribosyl-transferase (*FUR1*) (**Supplementary Table 6)**.

###### **Mating type identification**

In yeast, the mating-types (*MAT*) locus is a specialized region of the genome that regulates sexual identity, specifying cells as α, a, or α/a types, and most species are outcrossing with two opposite *MAT* locus^23^. To determine the mating-type-loci (*MTL*) of our *Candida auris* isolates, BLASTp v2.9.0^17^ searches were conducted on the 18 representative genomes using reference sequences from *MTLa* strain B8441 (TQR88726.1, TQR88727.1) and *MTLα* strain B11221 (TQB77621.1). BLAST searching results are in **Supplementary Table 5**.

###### **Characterization of Short Tandem Repeat (STR) loci**

All nine STR markers were located in the *Candida auris* B11205 genome (GCA_016772135.1) by BLASTn^24^. Raw read data were aligned against the B11205 genome with BWA-MEM v0.7.17.2, resulting BAM were filtered and visualized with JBrowse v1.16.11 as previously described^2^. Aligned reads of all Clade VI isolates were visually inspected to determine the copy number of each marker. Retrieved copy numbers were analyzed with BioNumerics v7.6.1 (Applied Maths NV, Sint-Martems-Latem, Belgium) by employing the unweighted pair group method with arithmetic mean averages (UPGMA) using the multistate categorical similarity coefficient.

##### **Genome rearrangement analysis**

To analyze unique genome rearrangement patterns for *Candida auris* clades (I-VI)^25^, genomes from Clade II to VI were aligned to Clade I genome using NUCmerv3.1^21^ (with --maxmatch --nosimplify) and dnadiff v1.3 was used for extracting unique alignment results (1coords file). For this analysis, Clade I genome CA-AM1 (GCA_014673535.1) was used as a reference as it has complete chromosomes. Translocation, inversion, and no alignment regions were identified based on the alignment results. Additionally, the centromere region of each chromosome was located by identifying the region with GC frequency lower than 0·42, similar to the threshold used in the previous study^25^. Chord diagrams for visualizing rearrangement events (with a minimum size of 10,000 nucleotides) were generated using pyCircos v0.3.0 (<https://github.com/ponnhide/pyCircos>).

##### **Phylogenetic timing analysis**

Bayesian phylogenies were generated using BEAST v1.10.4 as previously described^26^. Publicly available data used for molecular timing analysis were retrieved from the SRA database (**Supplementary Table 2**). In short, WGS read data were aligned against the *Candida auris* genome B11205 (GCA_016772135.1) using BWA-MEM v0.7.17.2 and BAM files were subsequently filtered. SNPs were detected with FreeBayes v1.1.0.46 and manually filtered. The filtered VCF file was converted to FASTA alignments with vcf2phylip v2.8 and was converted to an xml file with BEAUti v1.10.4. Random nucleotides totaling the *Candida auris* genome size minus the number of positions in the vcf file were inserted in the xml file. Tip dates were set to the year of collection or to 2015 with an uncertainty of 5 if the collection year is unknown. The previously established mutation rate of 1.8695e-5 substitutions per site per year was used^27^. The GTR model with gamma variation among sites was used as the substitution model with a strict molecular clock. A coalescent exponential growth model was selected as the Tree Prior. The length of chain was set to 50 million and was run using an unweighted pair-group method with an arithmetic mean (UPGMA) tree. A total of 46% was discarded as burn-in and a maximum clade credibility tree was made with TreeAnnotator v1.8.4 and visualized with FigTree v1.4.4.

#### **Machine-learning models for detecting potential new clade**

Bayesian logistic regression models were trained based on SNP distances and previously reported clade information to learn a threshold for predicting whether a pair of genomes are from the same clade. The threshold was then used to determine the relationships between genome pairs (edge) in a graph, which captured clusters (connected components) representing existing and potential new clades. In this analysis, 3,651 publicly available WGS and three WGS datasets were generated in this study (**Supplementary Appendix 3**), of which 1,132 (31%) had previously reported clade information were included. All WGS samples were grouped into 19 quarterly datasets based on their date of public release (2019 - June 2023). At each time point, a graph was generated, where nodes represent genomes and edges link between two genomes that were predicted to belong to the same clade. The number of clusters (connected components) present in the graph represents the total number of clades predicted to be present in the dataset. An overview of our machine learning approach for detecting potential *Candida auris* new clade is depicted in the Figure 1 below.

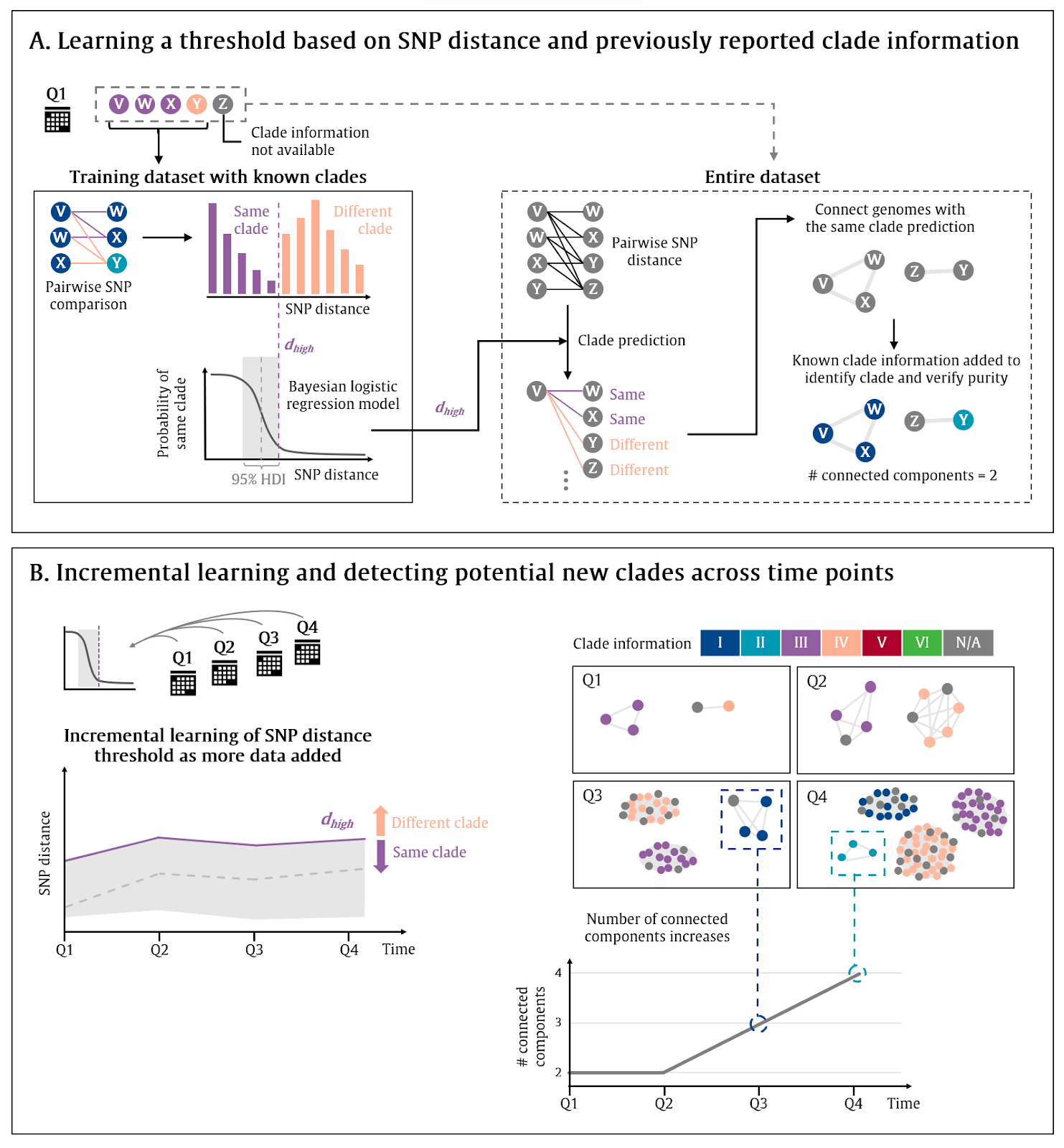

**Figure 1.** Overview of machine learning approach for detecting a potentially new *Candida auris* clade. **(A)** Learning a threshold based on SNP distance and previously reported clade information to predict whether a pair of genomes are from the same clade. **(B)** Incremental learning of the threshold and the detection of potentially new clades across time points.

##### **Overview of WGS data used for machine learning models for detecting potential new clade**

Our initial list of 4,475 *Candida auris* WGS accessions was obtained from Pathogen Detection (<https://www.ncbi.nlm.nih.gov/pathogens/>) and the previous study^1,2^, of which 1,285 have clade information available. Three additional Illumina WGS data of Clade VI isolates sequenced in this study were added to the set, making a total of 4,478 Illumina WGS accessions. Our three isolates and isolate SRR24877249 were then annotated as Clade VI.

Of the 4,775 WGS accessions, 4,344 were successfully analyzed, while 131 failed due to unable to download the sequence data (n=53), inadequate sequencing depth (<30✕; n=77), and unsuccessful genome assembly (likely because wrong sequencing files were uploaded; n=1). The following quality control parameters of the sequence data and assembled genomes were applied, and 3,654 WGS data, of which 1,132 (31%) had previously reported clade information, were used for the analysis (Supplementary Appendix 3).

1. Average sequencing depth is at least 30✕
2. Minimum read lengths of 100 nucleotides
3. Minimum read Phred score of 18
4. % of Q30 reads must be at least 80%
5. Assembled genome size is between 11,000,000-14,000,000 nucleotides
6. Assembled genome % GC content is between 43-47%
7. For species identification, *Candida auris* reference genome (GCF_003013715.1) was reported as the most similar genome with mash distance at most 0·04.

##### Bayesian logistic regression model

To predict whether a given pair of genomes belong to the same clade, Bayesian logistic regression models were trained based on SNP distance and previously reported clade information **(Figure 1A)**. Firstly, 6,674,031 SNP distance values among 3,654 isolates were calculated based on the Illumina variant calling results using B8441 (GCA_002759435.2) as a reference (343,270 unique SNPs detected). A large SNP distance matrix file is available upon request.

A Bayesian logistic regression model was defined as follows,

$$x= SNP distance$$

$$y= \{1: same clade, 0: different clades\}$$

$$\mu=\alpha+\beta x$$

$$\theta= sigmoid(\mu)$$

$$\hat{y}=Bernoulli(\theta, observed=y)$$

$$d=\frac{-\alpha}{\beta}$$

The prior of model parameters $\alpha$ and $\beta$ were set as a normal distribution $N(0, 10)$. The model was inferred using only the genome pairs with previously reported clade information using PyMC v3.11.5^28^ (draws=1000, cores=4, tune=1000). The decision boundary $d$ indicates the SNPs distance value $x$ where $\theta=0.5$, indicating the probability of 0.5 that a given pair of genomes are from the same clade. High-density interval ($d_{low}, d_{high}$) was calculated using arviz v0.12.1^29^, and a given genome pair with SNP distance $x\leq d_{high}$ was predicted to be from the same clade.

##### Generating a graph for detecting potential new clades

At each time point (in quarterly units; 19 quarters in total), a graph was generated, where nodes represent genomes and edges link between two genomes that were predicted to belong to the same clade **(Figure 1B)**. The numbers of connected components, which represent genome clusters, were counted using Networkx v3.1^30^. The numbers of genome clusters were tracked across the time points. The potential new clades could be flagged for further investigation when the number of connected components increases.

As the model was trained incrementally for each time point (in quarterly units), there was a possibility that some outlier might cause data drift, which could impact the threshold $d$. To allow users to monitor the model behavior, the distribution of threshold $d$ and a purity score $p$ was tracked across the time points (**Supplementary Table 7**). For each genome cluster $c$, the purity score $p_{c}=\frac{t_{c}}{(t_{c}+r_{c})}$, where $t_{c}$ is the number of genomes of the most frequent clade and $r_{c}$ is the number of genomes from the other clades, was calculated based on the genomes with previously reported clade information. The final purity score $p$ summarized the purity of the graph (weighted average based on the size of connected components), where $p=1$ suggests that every genome cluster consists of only one clade.

### **Supplementary Table 2**

| **ID** | **Country** | **Clade** | **SRA Accession Number** |
| --- | --- | --- | --- |
| B11205 | India | I | SRR3883436 |
| B11207 | India | I | SRR3883439 |
| L1537_2020 | Brazil | I | SRR13768964 |
| B8441 | Pakistan | I | SRR10851769 |
| B11112 | Pakistan | I | SRR3883473 |
| B11808 | South Korea | II | SRR10461263 |
| B14308 | USA | II | SRR10461147 |
| B13463 | Canada | II | SRR10461159 |
| B11229 | South Africa | III | SRR3883462 |
| B11230 | South Africa | III | SRR3883463 |
| B11225 | South Africa | III | SRR3883457 |
| B12037 | Canada | III | SRR10461253 |
| B12388 | USA | IV | SRR7909221 |
| B12177 | Venezuela | IV | SRR10461201 |
| B12336 | Colombia | IV | SRR7140028 |
| B12098 | Panama | IV | SRR10461248 |
| IFRC2087 | Iran | V | SRR9007776 |
| TMML616 | Iran | V | SRR18325431 |
| MRL40 | Iran | V | SRR18325430 |

**Supplementary Table 2.** Raw read data retrieved from the SRA database used for phylogenetic timing analysis (BEAST).

### **Supplementary Table 3**

| **Method** | **Well** | **Test** | **Mnemonic** | **Isolate A (F3485)** | **Isolate B (F1580)** | **Isolate C (F0083)** |
| --- | --- | --- | --- | --- | --- | --- |
| VITEK2 YST | 3 | L-Lysine-ARYLAMIDASE | LysA | - | - | - |
| VITEK2 YST | 4 | L-MALATE assimilation | IMLTa | + | + | (-) |
| VITEK2 YST | 5 | Leucine-ARYLAMIDASE | LeuA | + | + | + |
| VITEK2 YST | 7 | ARGININE | ARG | + | + | + |
| VITEK2 YST | 10 | ERYTHRITOL assimilation | ERYa | - | - | - |
| VITEK2 YST | 12 | GLYCEROL assimilation | GLYLa | - | - | - |
| VITEK2 YST | 13 | Tyrosine ARYLAMIDASE | TyrA | + | + | + |
| VITEK2 YST | 14 | BETA-N-ACETYL-GLUCOSAMINIDASE | BNAG | - | - | - |
| VITEK2 YST | 15 | ARBUTIN assimilation | ARBa | - | - | - |
| VITEK2 YST | 18 | AMYGDALIN assimilation | AMYa | - | - | - |
| VITEK2 YST | 19 | D-GALACTOSE assimilation | dGALa | - | - | - |
| VITEK2 YST | 20 | GENTOBIOSE assimilation | GENa | - | - | - |
| VITEK2 YST | 21 | D-GLUCOSE assimilation | dGLUa | + | + | + |
| VITEK2 YST | 23 | LACTOSE assimilation | LACa | - | - | - |
| VITEK2 YST | 24 | METHYL-A-D-GLUCOPYRANOSIDE assimilation | MAdGa | - | - | - |
| VITEK2 YST | 26 | D-CELLOBIOSE assimilation | dCELa | - | - | - |
| VITEK2 YST | 27 | GAMMA-GLUTAMYL-TRANSFERASE | GGT | - | - | - |
| VITEK2 YST | 28 | D-MALTOSE assimilation | dMALa | + | + | + |
| VITEK2 YST | 29 | D-RAFFINOSE assimilation | dRAFa | + | + | + |
| VITEK2 YST | 30 | PNP-N-acetyl-BD-galactosaminidase 1 | NAGA1 | (-) | - | - |
| VITEK2 YST | 32 | D-MANNOSE assimilation | dMNEa | + | + | + |
| VITEK2 YST | 33 | D-MELIBIOSE assimilation | dMELa | - | - | - |
| VITEK2 YST | 34 | D-MELEZITOSE assimilation | dMLZa | + | + | + |
| VITEK2 YST | 38 | L-SORBOSE assimilation | ISBEa | - | - | - |
| VITEK2 YST | 39 | L-RHAMNOSE assimilation | IRHAa | + | + | + |
| VITEK2 YST | 40 | XYLITOL assimilation | XLTa | + | - | + |
| VITEK2 YST | 42 | D-SORBITOL assimilation | dSORa | + | + | + |
| VITEK2 YST | 44 | SACCHAROSE/SUCROSE assimilation | SACa | + | + | + |
| VITEK2 YST | 45 | UREASE | URE | - | - | - |
| VITEK2 YST | 46 | ALPHA-GLUCOSIDASE | AGLU | + | + | + |
| VITEK2 YST | 47 | D-TURANOSE assimilation | dTURa | + | + | + |
| VITEK2 YST | 48 | D-TREHALOSE assimilation | dTREa | + | + | + |
| VITEK2 YST | 49 | NITRATE assimilation | NO3a | - | - | - |
| VITEK2 YST | 51 | L-ARABINOSE assimilation | IARAa | - | - | - |
| VITEK2 YST | 52 | D-GALACTURONATE assimilation | dGATa | + | + | + |
| VITEK2 YST | 53 | ESCULIN hydrolysis | ESC | - | - | - |
| VITEK2 YST | 54 | L-GLUTAMATE assimilation | IGLTa | + | + | + |
| VITEK2 YST | 55 | D-XYLOSE assimilation | dXYLa | - | - | - |
| VITEK2 YST | 56 | DL-LACTATE assimilation | LATa | - | - | - |
| VITEK2 YST | 58 | ACETATE assimilation | ACEa | + | + | + |
| VITEK2 YST | 59 | CITRATE (SODIUM) assimilation | CITa | + | + | + |
| VITEK2 YST | 60 | GLUCURONATE ASSIMILATION | GRTas | + | + | + |
| VITEK2 YST | 61 | L-PROLINE assimilation | IPROa | + | + | + |
| VITEK2 YST | 62 | 2-KETO-D-GLUCONATE assimilation | 2KGa | + | + | + |
| VITEK2 YST | 63 | N-ACETYL-GLUCOSAMINE assimilation | NAGa | + | + | + |
| VITEK2 YST | 64 | D-GLUCONATE assimilation | dGNTa | + | + | + |
| API20C AUX | 1 | None | 0 | - | - | - |
| API20C AUX | 2 | D-Glucose | GLU | + | + | + |
| API20C AUX | 3 | Glycerol | GLY | + | - | + |
| API20C AUX | 4 | Calcium 2-keto-gluconate | 2KG | + | + | + |
| API20C AUX | 5 | L-Arabinose | ARA | - | - | - |
| API20C AUX | 6 | D-Xylose | XYL | - | - | - |
| API20C AUX | 7 | Adonitol | ADO | - | - | - |
| API20C AUX | 8 | Xylitol | XLT | - | - | - |
| API20C AUX | 9 | D-Galactose | GAL | - | - | - |
| API20C AUX | 10 | Inositol | INO | - | - | - |
| API20C AUX | 11 | D-Sorbitol | SOR | + | + | + |
| API20C AUX | 12 | Methyl-αD-glucopyranoside | MDG | - | - | - |
| API20C AUX | 13 | N-Acetyl-glucosamine | NAG | + | + | + |
| API20C AUX | 14 | D-Cellobiose | CEL | - | - | - |
| API20C AUX | 15 | D-Lactose (bovine origin) | LAC | - | - | - |
| API20C AUX | 16 | D-Maltose | MAL | + | + | + |
| API20C AUX | 17 | D-Saccharose (sucrose) | SAC | + | + | + |
| API20C AUX | 18 | D-Trehalose | TRE | - | - | + |
| API20C AUX | 19 | D-Melezitose | MLZ | + | + | + |
| API20C AUX | 20 | D-Raffinose | RAF | + | + | + |
| API20C AUX | 21 | Hyphae/ Pseudophyphae | H/PH | - | - | - |
| API20C AUX | API Web | API 20 C AUX V5.0 Profile | Profile | 6102133 | 2102133 | 6102173 |
| MALDI-TOF | NA | MALDI Biotyper MSP Identification Standard Method 1.1 (Bruker) | NA | Candida auris (2.21) | Candida auris (2.27) | Candida auris (2.32) |
| MALDI-TOF | NA | CDC Micronet | NA | Candida auris (2.354) | Candida auris (2.232) | Candida auris (2.292) |

**Supplementary Table 3.** Phenotypic test results and MALDI identification scores for the Singapore Clade VI isolates.

### **Supplementary Figure 1**

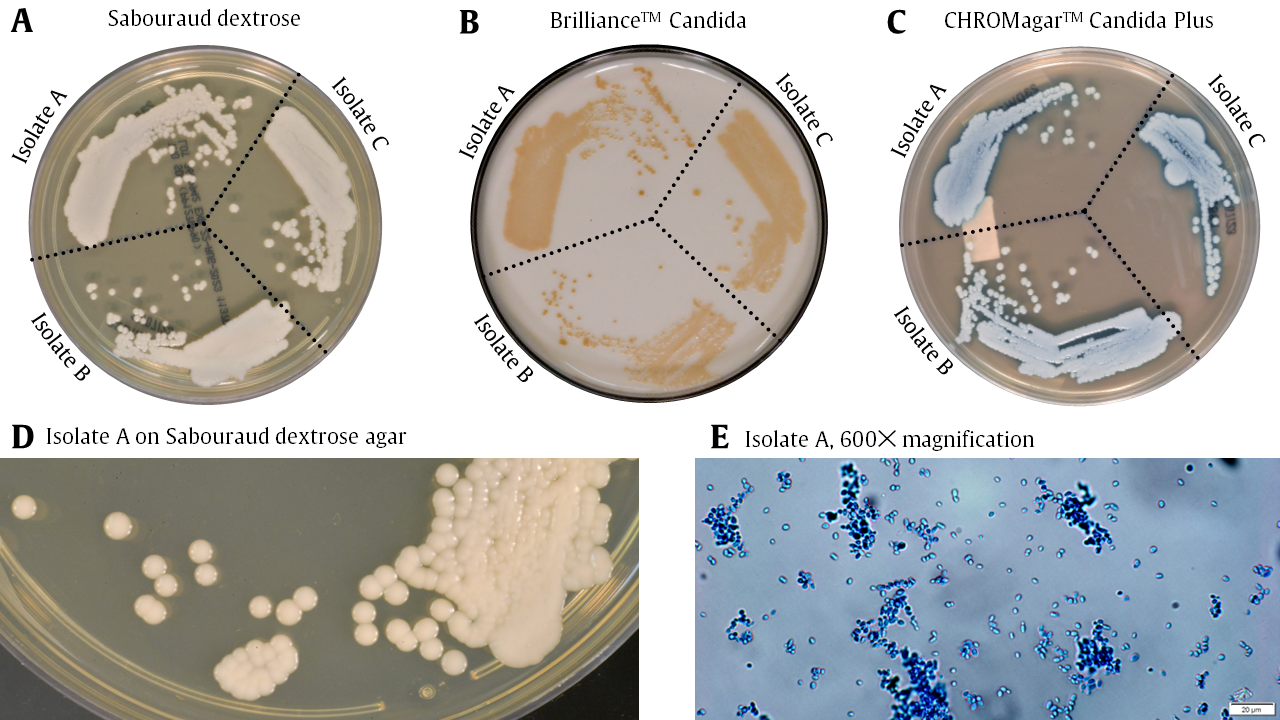

**Supplementary Figure 1.** Colony and microscopic morphology of Singapore Clade VI *Candida auris* isolates A (F3485), B (F1580), and C (F0083). (A) Colony morphology on Sabouraud dextrose agar (SDA) (Waltham, MA, United States, Thermo Fisher Scientific). (B) Colony morphology on Brilliance^TM^ Candida Agar (Oxoid, REF PO1034A, Basingstoke, United Kingdom, carried by Thermo Scientific^TM^). (C) Colony morphology on CHROMagar^TM^ Candida Plus agar (Paris, France, CHROMagar^TM^, REF CA242). (D) Close-up view of colony morphology on SDA of Isolate A (F3485). (E) Lactophenol Cotton Blue (Sigma Aldrich, St Louis, MO, United States) slide mount of Isolate A (F3485) at 600✕ magnification.

### **Supplementary Table 4**

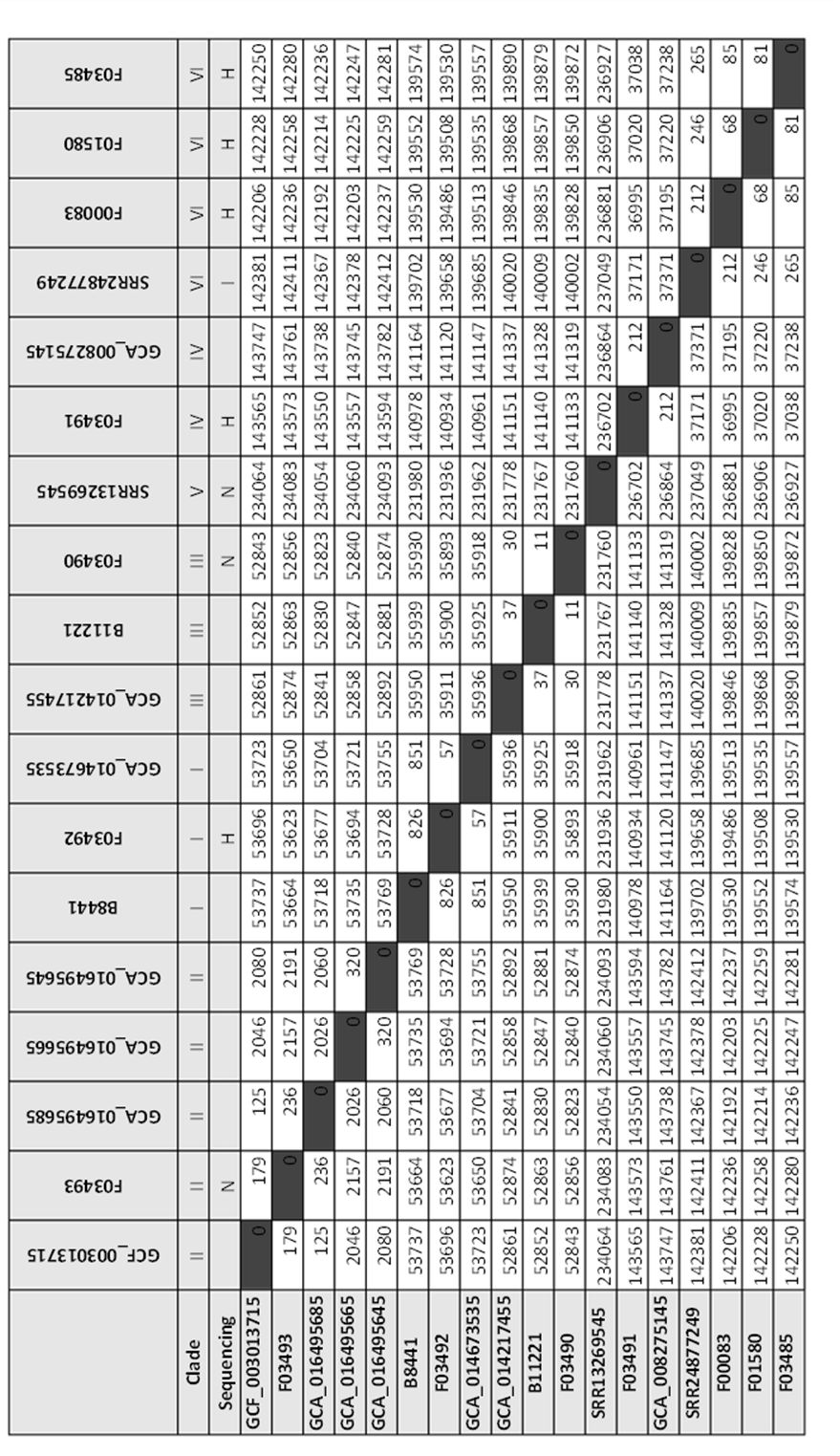

**Supplementary Table 4**. SNP matrix within clades. Abbreviations: N, nanopore sequencing. H, hybrid assemblies (Illumina and nanopore).

### **Supplementary Figure 2**

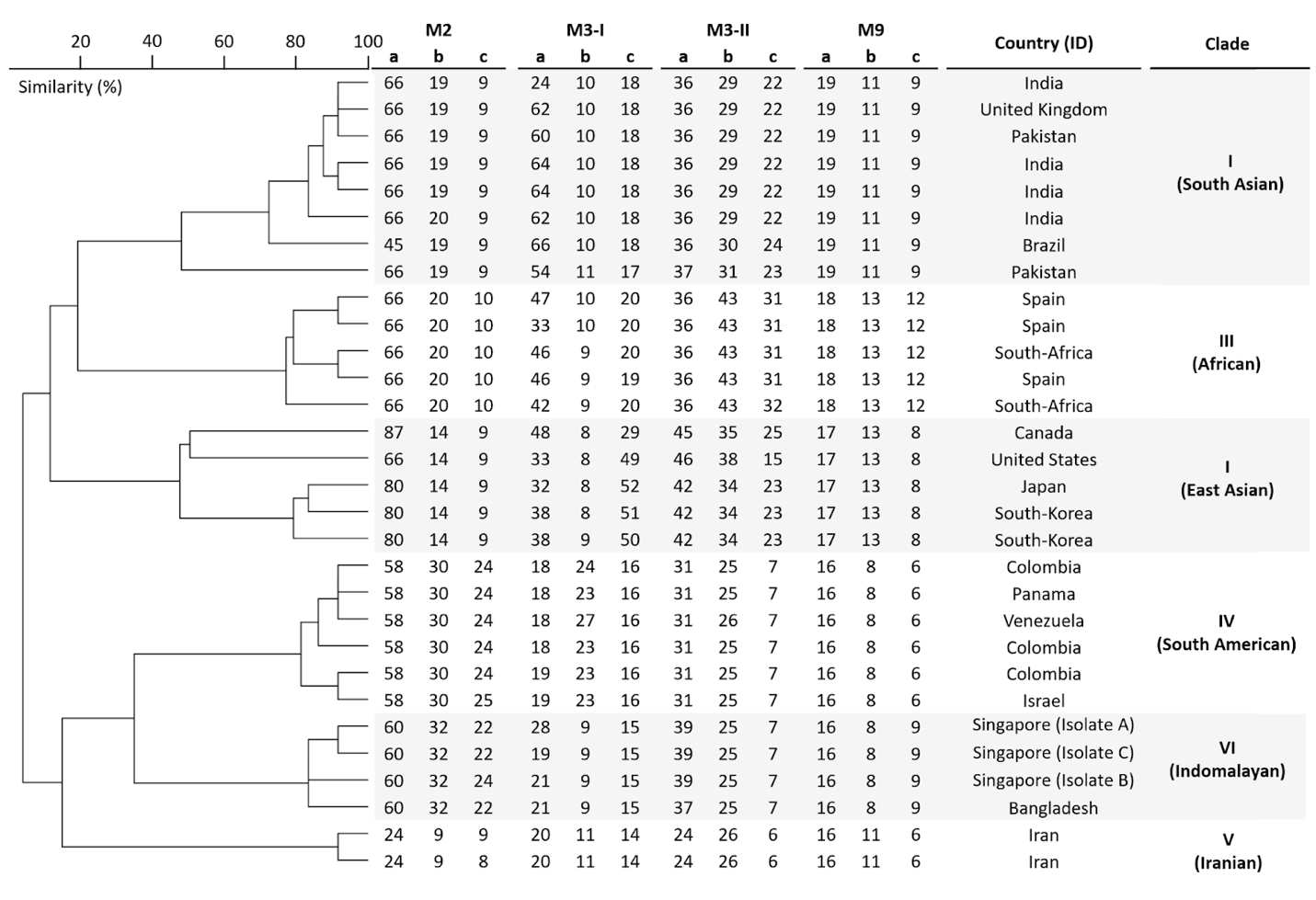

**Supplementary Figure 2.** *In silico* short tandem repeat (STR) analysis results. The dendrogram indicates the presence of six unique clusters based on STR profiles. Three Singaporean Clade VI isolates, A (F3485), B (F1580), and C (F0083), and one additional independently submitted whole-genome data from Bangladesh (SRR24877249), formed a distinct Clade VI cluster based on STR profiles.

### **Supplementary Figure 3**

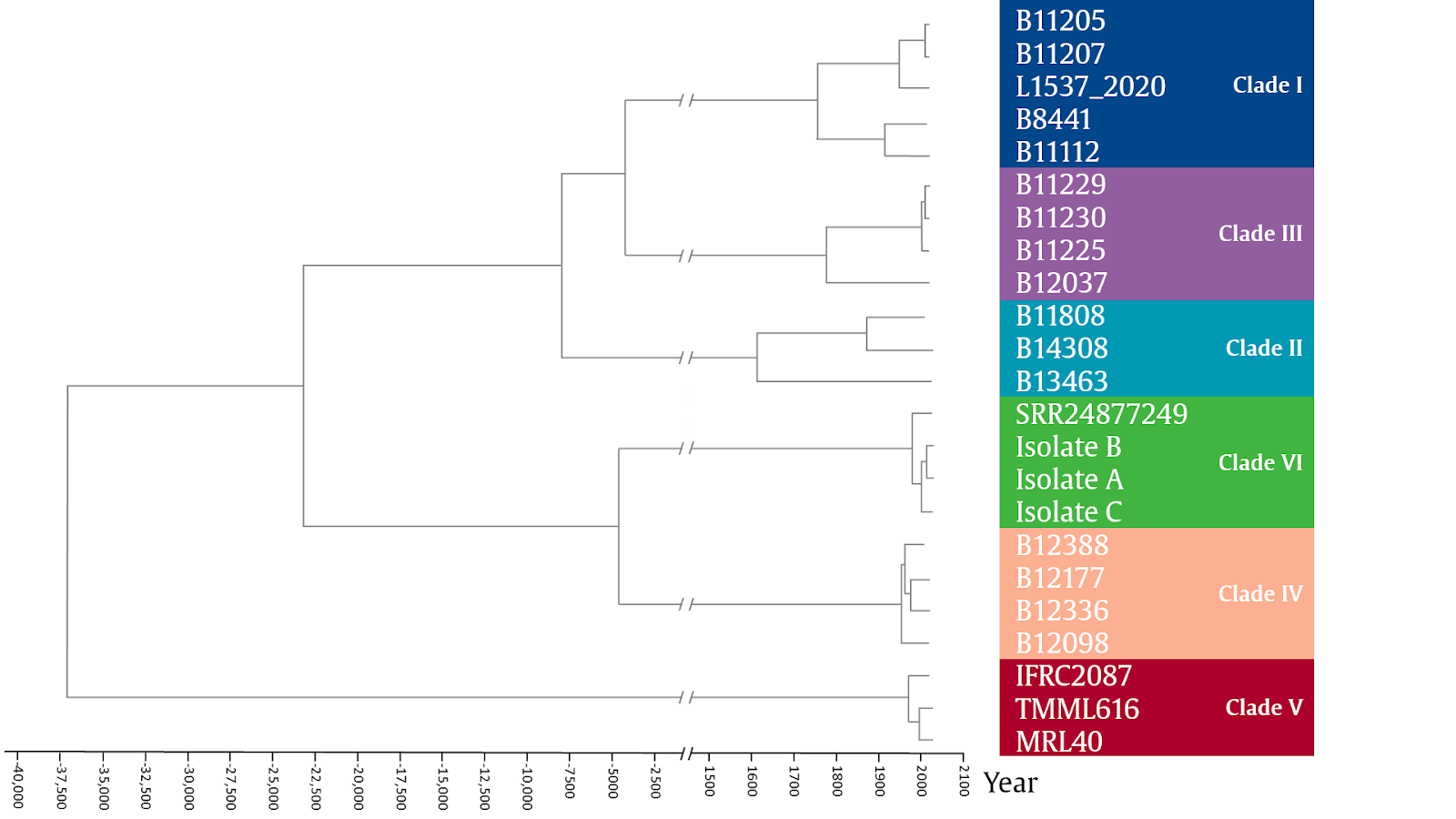

**Supplementary Figure 3.** Phylogenetic tree of six *Candida auris* clades with estimated dates of the most recent common ancestor. The tree is generated with BEAST v1.10.4 using a strict molecular clock with a coalescent model. Divergence times between selected isolates, collected between 2004 and 2023, were estimated with Bayesian inference. The GTR model was utilized as the substitution model with a coalescent exponential growth model. The estimated time to the most recent common ancestor (TMRCA) for clade VI occurred around 1989 (95% Highest probability density [HPD] 29.6 – 40.2 years ago). Clade VI and the most closely related clade IV had an estimated divergence at 4831 BCE (HPD 272.4 – 311.6 years ago). By including all reported six clades, the TMRCA for *Candida auris* as a species was set at 37,000 BCE approximately. Isolates sequenced and reported in this study: Isolate B (F1580, Patient B); Isolate A (F3485, Patient A); Isolate C (F0083, Patient C).

### **Supplementary Figure 4**

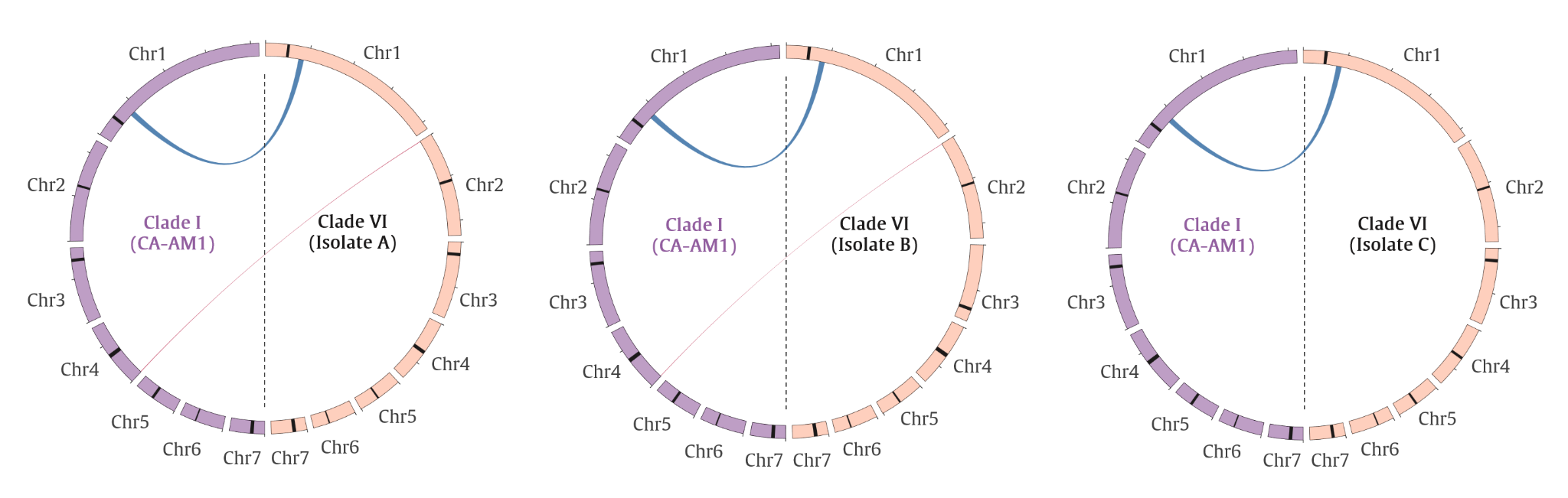

**Supplementary figure 3**. Chromosomal rearrangement in Clade VI *Candida auris* isolates. Chord diagram showing translocations and inversions observed in Isolates A (F3485), B (F1580), and C (F0083), using CA-AM1 (GCA_014673535) (Clade I) as a reference. Only rearrangement events of size >10,000bp are displayed.

### **Supplementary Table 5**

| **Query** | **Query name** | **Query WGS clade** | **Subject ID** | **Subject name** | **Subject length** | **Alignment subject coverage** | **Alignment identity** |
| --- | --- | --- | --- | --- | --- | --- | --- |
| B11221 | B11221 | III | gb\|TQB77621.1\| | MTLalpha1 | 172 | 95.93 | 99.39 |
| B8441 | B8441 | I | gb\|TQR88727.1\| | MTLa1 | 99 | 100.00 | 97.98 |
| B8441 | B8441 | I | gb\|TQR88726.1\| | MTLa2 | 158 | 51.27 | 100.00 |
| B8441 | B8441 | I | gb\|TQR88726.1\| | MTLa2 | 158 | 48.10 | 98.68 |
| F00083 | Isolate C | VI | gb\|TQB77621.1\| | MTLalpha1 | 172 | 97.67 | 95.24 |
| F01580 | Isolate B | VI | gb\|TQB77621.1\| | MTLalpha1 | 172 | 97.67 | 95.24 |
| F03485 | Isolate A | VI | gb\|TQB77621.1\| | MTLalpha1 | 172 | 97.67 | 95.24 |
| F03490 | ATCC5002 | III | gb\|TQB77621.1\| | MTLalpha1 | 172 | 95.93 | 99.39 |
| F03491 | ATCC5003 | IV | gb\|TQR88727.1\| | MTLa1 | 99 | 100.00 | 96.97 |
| F03491 | ATCC5003 | IV | gb\|TQR88726.1\| | MTLa2 | 158 | 51.27 | 97.53 |
| F03491 | ATCC5003 | IV | gb\|TQR88726.1\| | MTLa2 | 158 | 48.10 | 97.37 |
| F03492 | ATCC5000 | I | gb\|TQR88727.1\| | MTLa1 | 99 | 100.00 | 97.98 |
| F03492 | ATCC5000 | I | gb\|TQR88726.1\| | MTLa2 | 158 | 51.27 | 100.00 |
| F03492 | ATCC5000 | I | gb\|TQR88726.1\| | MTLa2 | 158 | 48.10 | 98.68 |
| F03493 | ATCC5001 | II | gb\|TQB77621.1\| | MTLalpha1 | 172 | 95.93 | 98.79 |
| GCA_008275145 | B11245 | IV | gb\|TQR88726.1\| | MTLa2 | 158 | 51.27 | 97.53 |
| GCA_008275145 | B11245 | IV | gb\|TQR88726.1\| | MTLa2 | 158 | 48.10 | 97.37 |
| GCA_014217455 | A1 | III | gb\|TQB77621.1\| | MTLalpha1 | 172 | 95.93 | 99.39 |
| GCA_014673535 | CA-AM1 | I | gb\|TQR88727.1\| | MTLa1 | 99 | 100.00 | 97.98 |
| GCA_014673535 | CA-AM1 | I | gb\|TQR88726.1\| | MTLa2 | 158 | 51.27 | 100.00 |
| GCA_014673535 | CA-AM1 | I | gb\|TQR88726.1\| | MTLa2 | 158 | 48.10 | 98.68 |
| GCA_016495645 | B12043 | II | gb\|TQB77621.1\| | MTLalpha1 | 172 | 95.93 | 98.79 |
| GCA_016495665 | B13463 | II | gb\|TQB77621.1\| | MTLalpha1 | 172 | 95.93 | 98.79 |
| GCA_016495685 | B11809 | II | gb\|TQB77621.1\| | MTLalpha1 | 172 | 95.93 | 98.79 |
| GCF_003013715 | B11220 | II | gb\|TQB77621.1\| | MTLalpha1 | 172 | 95.93 | 98.79 |
| SRR13269545 | SRR13269545 | V | gb\|TQR88727.1\| | MTLa1 | 99 | 100.00 | 97.98 |
| SRR13269545 | SRR13269545 | V | gb\|TQR88726.1\| | MTLa2 | 158 | 51.27 | 92.59 |
| SRR13269545 | SRR13269545 | V | gb\|TQR88726.1\| | MTLa2 | 158 | 48.10 | 94.74 |
| SRR24877249 | SRR24877249 | VI | gb\|TQB77621.1\| | MTLalpha1 | 172 | 97.67 | 95.24 |

**Supplementary Table 5.** Mating type loci analysis results.

### **Supplementary results and discussions**

##### **Additional results and discussions on antifungal susceptibility testing and antifungal resistance genes characterization**

Antifungal susceptibility testing of the Singapore Isolates A and B showed wild-type minimum inhibitory concentrations (MICs) for all tested antifungals^31^. Isolate C showed MIC of 2 mg/L for Amphotericin B, which is considered resistant based on the CDC tentative MIC breakpoint^32^. However, a recent report found that Sensititre YeastOne overestimates amphotericin B resistance, hence this finding of a higher amphotericin MIC in Isolate C needs to be interpreted with caution^33^. No known resistance-conferring mutations were found in the following genes: lanosterol 14 α-demethylase (*ERG11*), Sterol 24-C-methyltransferase (*ERG6*), 1,3-beta-glucan synthase (*FKS1*), zinc-cluster transcription factor-encoding gene *TAC1B*, and uracil phosphoribosyl-transferase (*FUR1*), as shown in **Supplementary Table 6** below.

###### **Supplementary Table 6**

Antifungal resistance mutations analysis results. Abbreviations: NA, not applicable.

**Table 6A**. Known azole resistance-conferring mutations in lanosterol 14 α-demethylase (*ERG11*)

| **Query_Label** | **F126L** | **K143R** | **Y132F** | **F444L** |
| --- | --- | --- | --- | --- |
| **GCA_016495645 \| Clade II** | NA | NA | NA | NA |
| **GCA_016495665 \| Clade II** | NA | NA | NA | NA |
| **GCA_016495685 \| Clade II** | NA | NA | NA | NA |
| **GCF_003013715 \| Clade II** | NA | NA | NA | NA |
| **F03493 \| Clade II** | NA | NA | NA | NA |
| **GCA_014217455 \| Clade III** | missense_variant | NA | NA | NA |
| **B11221 \| Clade III** | missense_variant | NA | NA | NA |
| **F03490 \| Clade III** | missense_variant | NA | NA | NA |
| **B8441 \| Clade I** | NA | NA | NA | NA |
| **GCA_014673535 \| Clade I** | NA | missense_variant | NA | NA |
| **F03492 \| Clade I** | NA | missense_variant | NA | NA |
| **SRR13269545 \| Clade V** | NA | NA | NA | NA |
| **F03491 \| Clade IV** | NA | NA | missense_variant | NA |
| **GCA_008275145 \| Clade IV** | NA | NA | missense_variant | NA |
| **F03485 \| Clade VI** | NA | NA | NA | NA |
| **F01580 \| Clade VI** | NA | NA | NA | NA |
| **SRR24877249 \| Clade VI** | NA | NA | NA | NA |
| **F00083 \| Clade VI** | NA | NA | NA | NA |

**Table 6B**. Known mutations in Sterol 24-C-methyltransferase (*ERG6*) associated with amphotericin B resistance

| **Query_Label** | **G403T** |
| --- | --- |
| **GCA_016495645 \| Clade II** | NA |
| **GCA_016495665 \| Clade II** | NA |
| **GCA_016495685 \| Clade II** | NA |
| **GCF_003013715 \| Clade II** | NA |
| **F03493 \| Clade II** | NA |
| **GCA_014217455 \| Clade III** | NA |
| **B11221 \| Clade III** | NA |
| **F03490 \| Clade III** | NA |
| **B8441 \| Clade I** | NA |
| **GCA_014673535 \| Clade I** | NA |
| **F03492 \| Clade I** | NA |
| **SRR13269545 \| Clade V** | NA |
| **F03491 \| Clade IV** | NA |
| **GCA_008275145 \| Clade IV** | NA |
| **F03485 \| Clade VI** | NA |
| **F01580 \| Clade VI** | NA |
| **SRR24877249 \| Clade VI** | NA |
| **F00083 \| Clade VI** | NA |

**Table 6C**. Known echinocandin resistance-conferring mutations in 1,3-beta-glucan synthase (*FKS1*)

| **Query_Label** | **S639Y/P/F** |
| --- | --- |
| **GCA_016495645 \| Clade II** | NA |
| **GCA_016495665 \| Clade II** | NA |
| **GCA_016495685 \| Clade II** | NA |
| **GCF_003013715 \| Clade II** | NA |
| **F03493 \| Clade II** | NA |
| **GCA_014217455 \| Clade III** | NA |
| **B11221 \| Clade III** | NA |
| **F03490 \| Clade III** | NA |
| **B8441 \| Clade I** | NA |
| **GCA_014673535 \| Clade I** | NA |
| **F03492 \| Clade I** | NA |
| **SRR13269545 \| Clade V** | NA |
| **F03491 \| Clade IV** | NA |
| **GCA_008275145 \| Clade IV** | NA |
| **F03485 \| Clade VI** | NA |
| **F01580 \| Clade VI** | NA |
| **SRR24877249 \| Clade VI** | NA |
| **F00083 \| Clade VI** | NA |

**Table 6D**. Known mutations in zinc-cluster transcription factor-encoding gene (*TAC1B)* associated with azole resistance

| **Query_Label** | **S611P** | **A640V** |
| --- | --- | --- |
| **GCA_016495645 \| Clade II** | NA | NA |
| **GCA_016495665 \| Clade II** | NA | NA |
| **GCA_016495685 \| Clade II** | NA | NA |
| **GCF_003013715 \| Clade II** | NA | NA |
| **F03493 \| Clade II** | NA | NA |
| **GCA_014217455 \| Clade III** | NA | NA |
| **B11221 \| Clade III** | NA | NA |
| **F03490 \| Clade III** | NA | NA |
| **B8441 \| Clade I** | NA | NA |
| **GCA_014673535 \| Clade I** | NA | missense_variant |
| **F03492 \| Clade I** | NA | missense_variant |
| **SRR13269545 \| Clade V** | NA | NA |
| **F03491 \| Clade IV** | NA | NA |
| **GCA_008275145 \| Clade IV** | NA | NA |
| **Isolate A (F3485) \| Clade VI** | NA | NA |
| **Isolate B (F1580) \| Clade VI** | NA | NA |
| **SRR24877249 \| Clade VI** | NA | NA |
| **Isolate C (F0083) \| Clade VI** | NA | NA |

**Table 6E**. Known mutations in uracil phosphoribosyl-transferase (*FUR1*) associated with flucytosine resistance.

| **Query_Label** | **F211I** |
| --- | --- |
| **GCA_016495645 \| Clade II** | NA |
| **GCA_016495665 \| Clade II** | NA |
| **GCA_016495685 \| Clade II** | NA |
| **GCF_003013715 \| Clade II** | NA |
| **F03493 \| Clade II** | NA |
| **GCA_014217455 \| Clade III** | NA |
| **B11221 \| Clade III** | NA |
| **F03490 \| Clade III** | NA |
| **B8441 \| Clade I** | NA |
| **GCA_014673535 \| Clade I** | NA |
| **F03492 \| Clade I** | NA |
| **SRR13269545 \| Clade V** | NA |
| **F03491 \| Clade IV** | NA |
| **GCA_008275145 \| Clade IV** | NA |
| **F03485 \| Clade VI** | NA |
| **F01580 \| Clade VI** | NA |
| **SRR24877249 \| Clade VI** | NA |
| **F00083 \| Clade VI** | NA |

##### **Additional discussion on divergence time**

With a molecular clock analysis, we estimated the TMRCA of Clade VI in addition to the divergence between clades. Bayesian phylogenies estimated a recent TMRCA of Clade VI around 1989, suggesting a diversification of the included clade VI isolates before the detection of this clade and a recent expansion into the human population. Divergence times of Clades IV and V were found to be highly similar to Clade VI, and all occurred at the end of the 19th century. The TMRCA of the most closely related Clade IV was set around 4600 BCE suggesting an old separation of these two clades. Estimated TMRCAs of Clades I to IV are similar as previously described^27^ and are supported by the genetic diversity found in the current study. Speciation of different *Candida auris* clades is linked to the geographic separation. However, the split between Clade VI and IV cannot be explained by the continental drift dispersal hypothesis as proposed for *Cryptococcus neoformans* and *gatti* species complexes^34^. Many scientists believe *Candida auris* occupies a special ecological niche^34^. It is probable that the ancestor of Clade VI was introduced into tropical Asia through migratory birds or oceanic currents.

##### **Additional discussion on mating type**

*Candida* species are successful yeast pathogens originally thought to be asexual, but several are now recognized as sexual or parasexual^23,35^. *Candida auris* is haploid, and although both mating types (α and a) have been reported in different clades, there is no formal report of sexual reproduction observed in *Candida auris* since different clades were largely geographically isolated ^1,36^. Most *Candida auris* isolates in Singapore belonged to Clade I which carried *MTLa* allele^37^ with sporadic isolates belonging to Clade II carrying *MTLα* allele. The presence of Clade VI in Singapore and in the Indomalayan region suggests that *Candida auris* clades carrying opposite mating types may be co-circulating. In the Southeast Asia and South Asia context, the coexistence of mixed distributions of the widely circulating Clade I carrying *MTLa* allele, and Clade VI carrying *MTLα* allele may theoretically provide opportunities for mating and recombination among strains from different clades to generate genetically diverse progenies^1,38^.

##### **Additional discussion on the detection and containment of *Candia auris* in Singapore**

As a regional hub, Singapore caters to a large international visitor volume annually that exceeds its resident population size. As such, the importation and dissemination of pathogens, and their associated antimicrobial resistance, is a constant public health threat to Singapore. However, this unique confluence also positions Singapore as an ideal sentinel site for identifying emergent public health threats. In the case of *Candida auris*, Singaporean authorities have taken a proactive stance and have put in place active surveillance programs and enhanced infection control measures in acute care hospitals since 2020. The index case presented in this study, Patient A, was found to be colonized as a result of the implementation of the active *Candida auris* surveillance program. Furthermore, none of the inpatient contacts of the three patients harboring Clade VI isolates tested positive for *Candida auris*, demonstrating the effectiveness of the infection prevention measures in place.

##### **Additional discussion on machine learning approach for new clade detection**

| **Time point** | **Number of nodes** | **Number of connected components** | **SNP distance threshold (mode)** | **SNP distance threshold (d_low_)** | **SNP distance threshold (d_high_)** | **Purity score** |
| --- | --- | --- | --- | --- | --- | --- |
| 2018 Q1 | 121 | 4 | 1499 | 1232 | 13488 | 1 |
| 2018 Q2 | 201 | 4 | 6654 | 5182 | 20947 | 1 |
| 2018 Q3 | 484 | 4 | 7691 | 6412 | 20848 | 1 |
| 2018 Q4 | 554 | 4 | 7925 | 6109 | 19469 | 1 |
| 2019 Q1 | 554 | 4 | 8064 | 5904 | 19717 | 1 |
| 2019 Q2 | 555 | 4 | 8281 | 6163 | 20571 | 1 |
| 2019 Q3 | 564 | 5 | 9617 | 7301 | 21237 | 1 |
| 2019 Q4 | 571 | 5 | 10702 | 8247 | 21040 | 1 |
| 2020 Q1 | 723 | 5 | 13937 | 11002 | 22498 | 1 |
| 2020 Q2 | 759 | 5 | 12278 | 10148 | 21980 | 1 |
| 2020 Q3 | 909 | 5 | 15605 | 10929 | 22844 | 1 |
| 2020 Q4 | 937 | 5 | 13372 | 10662 | 22037 | 1 |
| 2021 Q1 | 944 | 5 | 14183 | 10816 | 22020 | 1 |
| 2021 Q2 | 983 | 5 | 13532 | 11053 | 22014 | 1 |
| 2021 Q3 | 1094 | 5 | 12557 | 9762 | 20763 | 1 |
| 2021 Q4 | 1096 | 5 | 14848 | 11531 | 21973 | 1 |
| 2022 Q1 | 1171 | 5 | 14082 | 10370 | 21522 | 1 |
| 2022 Q2 | 1343 | 5 | 13908 | 10716 | 22219 | 1 |
| 2022 Q3 | 1771 | 5 | 13811 | 11164 | 22073 | 1 |
| 2022 Q4 | 1985 | 5 | 12646 | 10447 | 21387 | 1 |
| 2023 Q1 | 3219 | 5 | 13489 | 11254 | 21762 | 1 |
| 2023 Q2 | 3651 | 6 | 13745 | 10494 | 21594 | 1 |
| 2023 Q3 | 3654 | 6 | 11818 | 10481 | 21554 | 1 |

**Supplementary Table 7**. Model parameters learned across time points.

In this study, we demonstrated as a proof-of-concept that incorporation of machine learning approaches can automatically detect new *Candida auris* clades using whole-genome sequence data. These approaches can potentially enhance genomic surveillance by the early identification and investigation of outlier genomes. Here we discuss some challenges and limitations of this approach. Firstly, stringent quality control of input data is required. This resulted in significant data loss, which limits our ability to detect outlier genomes. Secondly, the model has limitations. Since only one reference genome was used to identify SNPs, there may be inherent biases and systematic errors in the resultant data^39^. To avoid the reliance on single reference genome, identifying SNPs based on pairwise genome alignment is an alternative way to measure genome relatedness measurement^21^, but this approach demands high computation resources for assembling the genomes. Techniques for detecting SNPs directly from two read files without requiring assembled genomes would require less computation resources^40^ and might also be applicable for detecting the outlier genomes. Thirdly, as the model learns and evolves with incremental data, the stability of the model needs to be monitored. This is mitigated by the implementation of various matrices to monitor model performance fluctuations, to allow the user-in-the-loop to detect suboptimal performance and therefore interpret the data with caution. Lastly, the model was designed to flag and alert potentially novel clade(s), which still required the flagged genomes to be investigated manually by the users.

This approach, however, allowed us to leverage all available *Candida auris* data at a given time point to arrive at a decision threshold, which is unlike the current approach of phylogenetic analysis, where a relatively small number of genomes need to be visualized in a tree structure. With a dynamic and data-driven threshold in place, algorithmic decisions can be made to flag outlier genomes automatically. This automatic surveillance for outlier genomes has the potential to augment microbial genomic surveillance and provide early alerts for potential public health threats.

31	Clinical and Laboratory Standard Institutes. Clinical and Laboratory Standard Institutes (CLSI). Epidemiological Cutoff Values for Antifungal Susceptibility Testing, 4th Edition. CLSI Supplement M57S. USA,2022. USA: Clinical and Laboratory Standard Institutes.

32	Antifungal Susceptibility Testing and Interpretation. 2023; published online April 5. <https://www.cdc.gov/fungal/candida-auris/c-auris-antifungal.html> (accessed July 27, 2023).

33	Siopi M, Peroukidou I, Beredaki M-I, *et al.* Overestimation of Amphotericin B Resistance in *Candida auris* with Sensititre YeastOne Antifungal Susceptibility Testing: a Need for Adjustment for Correct Interpretation. *Microbiol Spectr* 2023; **11**: e0443122.

34	Casadevall A, Freij JB, Hann-Soden C, Taylor J. Continental Drift and Speciation of the *Cryptococcus* *neoformans* and *Cryptococcus gattii* Species Complexes. *mSphere* 2017; **2**. DOI:[10.1128/mSphere.00103-17](http://dx.doi.org/10.1128/mSphere.00103-17).

35	Lee SC, Ni M, Li W, Shertz C, Heitman J. The evolution of sex: a perspective from the fungal kingdom. *Microbiol Mol Biol Rev* 2010; **74**: 298–340.

36	Muñoz JF, Gade L, Chow NA, *et al.* Genomic insights into multidrug-resistance, mating and virulence in *Candida auris* and related emerging species. *Nat Commun* 2018; **9**: 5346.

37	Tan YE, Teo JQ-M, Rahman NBA, *et al.* Candida auris in Singapore: Genomic epidemiology, antifungal drug resistance, and identification using the updated 8.01 VITEKⓇ2 system. *Int J Antimicrob Agents* 2019; **54**: 709–15.

38	Massic L, Gorzalski A, Siao DD, *et al.* Detection of five instances of dual-clade infections of *Candida auris* with opposite mating types in southern Nevada, USA. *Lancet Infect Dis* 2023; published online July 18. DOI:[10.1016/S1473-3099(23)00434-6](http://dx.doi.org/10.1016/S1473-3099(23)00434-6).

39	Valiente-Mullor C, Beamud B, Ansari I, *et al.* One is not enough: On the effects of reference genome for the mapping and subsequent analyses of short-reads. *PLoS Comput Biol* 2021; **17**: e1008678.

40	Gardner SN, Slezak T, Hall BG. kSNP3.0: SNP detection and phylogenetic analysis of genomes without genome alignment or reference genome. *Bioinformatics* 2015; **31**: 2877–8.
